# Strengthening vaccination delivery system resilience in the context of protracted humanitarian crisis: a realist-informed systematic review

**DOI:** 10.1101/2022.04.21.22273340

**Authors:** Sharif A. Ismail, Sze Tung Lam, Sadie Bell, Fouad M. Fouad, Karl Blanchet, Josephine Borghi

## Abstract

**Background:** Childhood vaccination is among the most effective public health interventions available for the prevention of communicable disease, but coverage in many humanitarian settings is sub-optimal. This systematic review critically evaluated peer-review and grey literature evidence on the effectiveness of system-level interventions for improving vaccination coverage in protracted crises, focusing on how they work, and for whom, to better inform preparedness and response for future crises.

**Methods:** Realist-informed systematic review of peer-reviewed and grey literature. Keyword-structured searches were performed in MEDLINE, EMBASE and Global Health, CINAHL, the Cochrane Collaboration and WHOLIS, and grey literature searches performed through the websites of UNICEF, the Global Polio Eradication Initiative (GPEI) and Technical Network for Strengthening Immunization Services. Results were independently double-screened for inclusion on title and abstract, and full text. Data were extracted using a pre-developed template, capturing information on the operating contexts in which interventions were implemented, intervention mechanisms, and vaccination-related outcomes. Study quality was assessed using the MMAT tool. Findings were narratively synthesised.

**Results:** 50 studies were included, most describing interventions applied in conflict or near-post conflict settings in sub-Saharan Africa, and complex humanitarian emergencies. Vaccination campaigns were the most commonly addressed adaptive mechanism (n=17). Almost all campaigns operated using multi-modal approaches combining service delivery through multiple pathways (fixed and roving), health worker recruitment and training and community engagement to address both vaccination supply and demand. Creation of collaterals through service integration showed generally positive evidence of impact on routine vaccination uptake by bringing services closer to target populations and leveraging trust that had already been built with communities. Robust community engagement emerged as a key unifying mechanism for outcome improvement across almost all of the intervention classes, in building awareness and trust among crisis-affected populations. Some potentially transformative mechanisms for strengthening resilience in vaccination delivery were identified, but evidence for these remains limited.

**Conclusion:** A number of interventions to support adaptations to routine immunisation delivery in the face of protracted crisis are identifiable, as are key unifying mechanisms (multi-level community engagement) apparently irrespective of context, but evidence remains piecemeal. Adapting these approaches for local system resilience-building remains a key challenge.

## INTRODUCTION

Childhood vaccination one of the most effective interventions in the armoury available to public health policymakers and practitioners (1–5), but there are major impediments to effective delivery in humanitarian settings and vaccination coverage in many of these contexts is low (6,7). Vaccine-preventable diseases (VPDs) have historically been, and continue to be, a major cause of mortality and morbidity population-wide, can undermine health service capacity through health worker absence due to illness, and lead to long-term reductions in economic productivity (5,8–10). In humanitarian settings, risks are intensified both to displaced populations (to whom vaccination delivery is often disrupted and in whom the prevalence of important risk factors for poor outcomes, such as malnutrition, tend to be higher than for settled populations) and to host communities (through disruptions to vaccination programme and indirectly through risks to herd protective effects).

The challenge of low coverage in humanitarian settings is compounded by a long-term shift in displacement patterns in humanitarian crises away from camps towards informal, urban or peri-urban settlements in which over 80% of refugees globally now reside (11–13). Populations living in these areas are often more mobile than those in camps, posing challenges for service delivery and for health information systems, especially so because many vaccines require multiple doses to ensure adequate protection. They also experience more pronounced barriers to care access through both national systems (in countries in which refugees and host communities are served through common public service pathways), and agency-led delivery systems (in countries where refugees continue to be served primarily through parallel arrangements). The evidence base to address this in crisis-affected settings is piecemeal (14–16). Most guidance focuses on acute rather than protracted crises (17,18), with little or no consideration of resilience-building or promoting measures that might improve national system resilience over the long-term.

The aim of this review was to identify system-level interventions to strengthen health system capacity to maintain and improve vaccination coverage in protracted humanitarian settings by bolstering the resilience of delivery systems. We use the definition of system resilience employed by Blanchet *et al* in their work on resilience governance, namely “the capacity of a health system to absorb, adapt or transform when exposed to a shock…and still retain the same control over its structure and functions” (19). This approach sets out three main mechanisms for system resilience: absorption, involving delivery of services at the same level (in terms of quantity, quality and equity) and using the same resources and capacities; adaptation, in which services are delivered at the same level but with fewer and/or different resources; and transformation, in which health system actors transform the structure and function of the system to respond to environmental change (19). The term “humanitarian crisis” also encapsulates a very wide range of contexts in which the pressures experienced by health systems, and the spectrum of appropriate responses to these, is considerable. Our analytical approach therefore drew on an emerging body of realist review work focusing not just on identifying which interventions work, but more particularly explaining *how* they work, and contextual factors shaping this (20–22), with a view to tailoring findings to different settings.

We developed a guiding conceptual framework that linked important health system functions (encapsulated by the health system building blocks) and aspects of the wider operating context, to mechanisms, and finally vaccination delivery-related outcomes, as illustrated in Figure 1. Given the diversity of intervention types considered here, developing a unitary programme theory (in line with convention for realist reviews) was neither feasible nor desirable. Instead, our focus was on describing the putative mechanisms by which included interventions worked, as an adjunct to a systematic review methodology (23–25). The process by which the conceptual framework was derived is outlined in more detail in **Appendix 1**.

**Figure 1.**
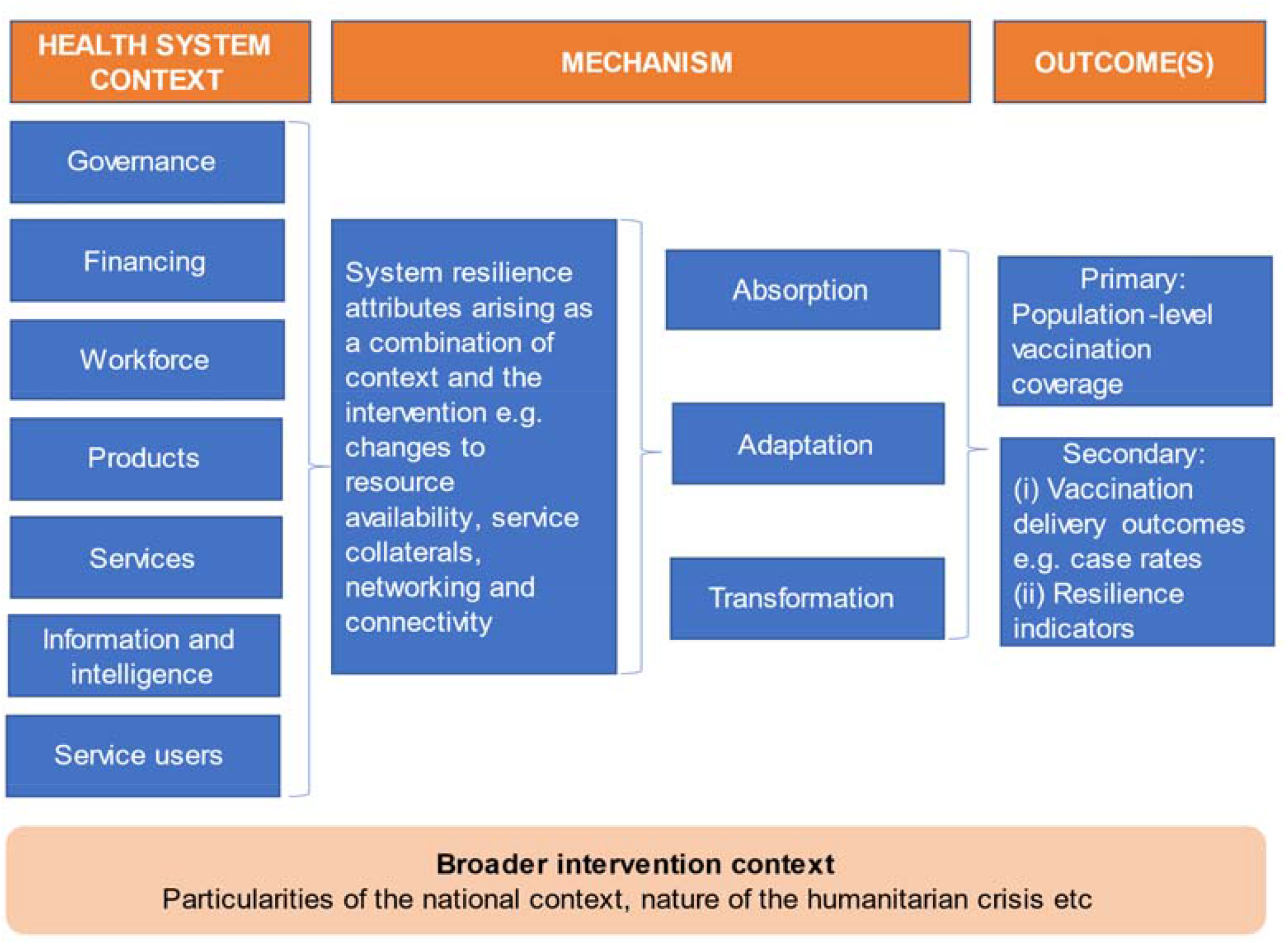
Guiding conceptual framework for the review, linking contextual aspects (conceptualised using the WHO building blocks framework, but also conditions linked to the broader national context and aspects of the specific humanitarian crisis in the setting for each intervention) and activities strengthening resilience attributes, the mechanisms for enhancing system resilience (absorption, adaptation, transformation) and finally the vaccination-related outcomes achieved (in this case population-level vaccination coverage for antigens included in the review). For more detail on how this framework was developed, please refer to **Appendix 1**.

## METHODS

This was a realist-informed systematic review of peer-reviewed and grey literature. The review was performed in accordance with the Preferred Reporting Items for Systematic Reviews and Meta-Analyses (PRISMA) guidelines (PROSPERO protocol reference CRD42021273124 – available <u>here</u>). We considered evidence relating to vaccination delivery for refugee, internally displaced and host community populations in LMICs affected by protracted humanitarian crises with a particular focus on children aged 0-5 as the target population for most routine vaccination programmes, but incorporating older displaced children, teenagers and adults in consideration of, for example, catch-up programmes.

### Definitions, inclusion and exclusion criteria

There is no broadly agreed definition of the term “protracted crisis” in the literature. We adopted the approach used in the Global Humanitarian Assistance Reports, including any country subject to at least five, consecutive years of UN-coordinated humanitarian action at any point between 2001 and 2021 (26). A shortlist of eligible countries was generated using this approach to determine which contexts to include in searches (see **Appendix 2**).

We gathered evidence on system strengthening interventions aimed at one or more of the WHO health system building blocks at meso- or macro-level, with explicit or potential effects on either or both of the primary and secondary outcome domains set out below. For inclusion, [i] there needed to be sufficient programmatic detail in the article to form a clear view of intervention design and how it had been implemented, and [ii] interventions had to operate at meso- or macro-level. Meso-level interventions included supply-side measures such as area-based interventions (i.e. district, governorate or equivalent level and above), health sector interventions addressing system resilience directly, or alternatively specifying activities under one or more of the WHO’s six health system building blocks; or interventions targeting specific tranches of the vaccination delivery pathway e.g. cold-chain maintenance; or demand-side interventions focused on refugee or otherwise displaced populations in crises with demonstrated effects on population health outcomes (e.g. cash transfer programmes), with the overall objective of increasing population demand for, and uptake of, vaccination. Macro-level interventions, on the other hand, addressed system resilience at national level directly, or concurrently addressed a number of the WHO’s six health system building blocks at national level; or described interventions targeting specific tranches of the vaccination delivery pathway at national level. Micro-level interventions such as the use of tailored text-messaging or other forms of individualised outreach were excluded from the review. Shorter-term activities such as vaccination campaigns were included provided there was demonstrable evidence that these contributed in some way to longer-term resilience-promotion (e.g. through training of workforce cadres, leveraging of existing structures in new ways). A full outline of inclusion and exclusion criteria is given in Table 1.

**Table 1.**
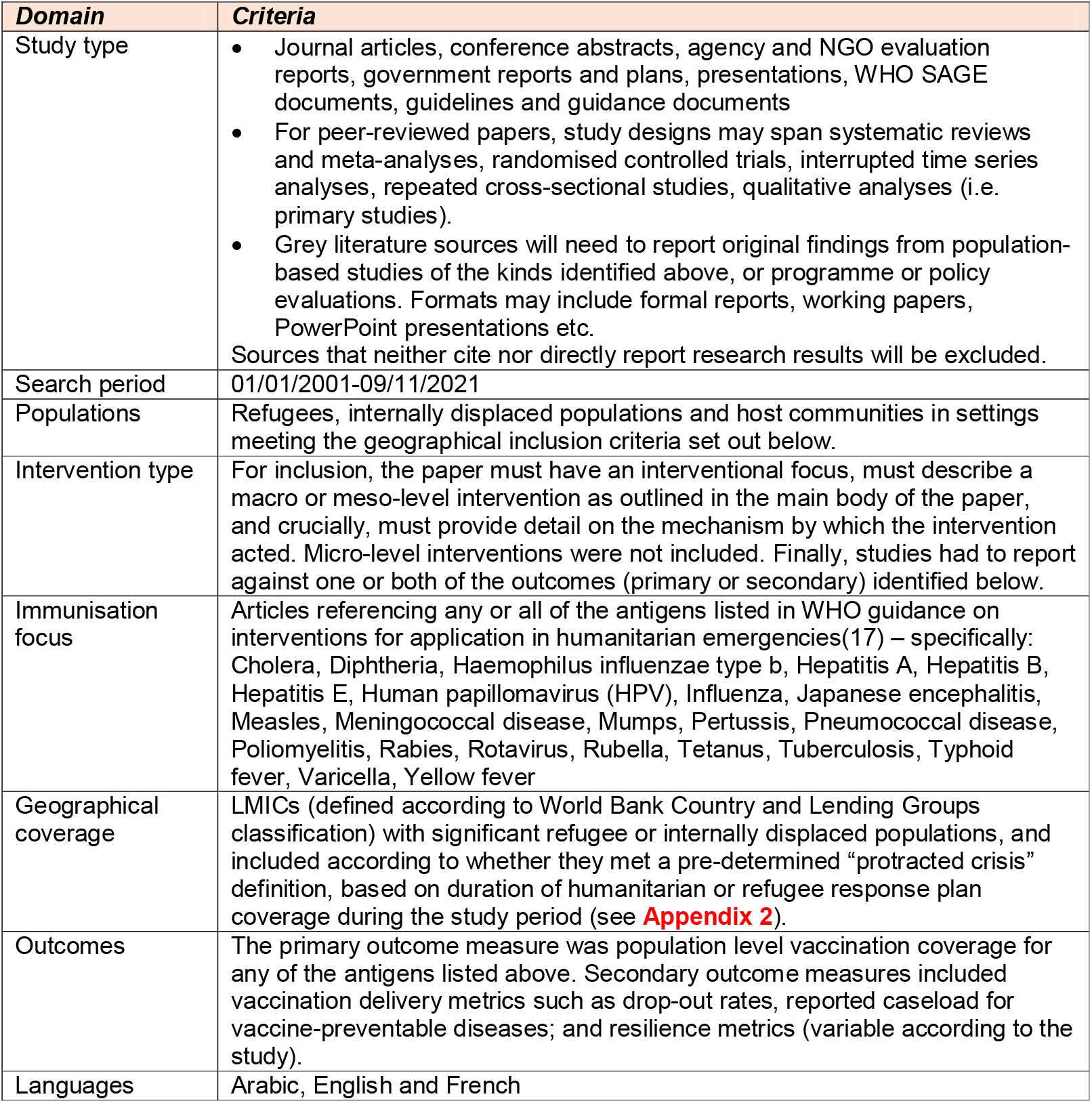
Inclusion criteria applied in the selection of studies for this paper.

| **Domain** | **Criteria** |
| --- | --- |
| Study type | Journal articles, conference abstracts, agency and NGO evaluation reports, government reports and plans, presentations, WHO SAGE documents, guidelines and guidance documents<br>For peer-reviewed papers, study designs may span systematic reviews and meta-analyses, randomised controlled trials, interrupted time series analyses, repeated cross-sectional studies, qualitative analyses (i.e. primary studies).<br>Grey literature sources will need to report original findings from population-based studies of the kinds identified above, or programme or policy evaluations. Formats may include formal reports, working papers, PowerPoint presentations etc.<br>Sources that neither cite nor directly report research results will be excluded. |
| Search period | 01/01/2001-09/11/2021 |
| Populations | Refugees, internally displaced populations and host communities in settings meeting the geographical inclusion criteria set out below. |
| Intervention type | For inclusion, the paper must have an interventional focus, must describe a macro or meso-level intervention as outlined in the main body of the paper, and crucially, must provide detail on the mechanism by which the intervention acted. Micro-level interventions were not included. Finally, studies had to report against one or both of the outcomes (primary or secondary) identified below. |
| Immunisation focus | Articles referencing any or all of the antigens listed in WHO guidance on interventions for application in humanitarian emergencies(17) – specifically: Cholera, Diphtheria, Haemophilus influenzae type b, Hepatitis A, Hepatitis B, Hepatitis E, Human papillomavirus (HPV), Influenza, Japanese encephalitis, Measles, Meningococcal disease, Mumps, Pertussis, Pneumococcal disease, Poliomyelitis, Rabies, Rotavirus, Rubella, Tetanus, Tuberculosis, Typhoid fever, Varicella, Yellow fever |
| Geographical coverage | LMICs (defined according to World Bank Country and Lending Groups classification) with significant refugee or internally displaced populations, and included according to whether they met a pre-determined “protracted crisis” definition, based on duration of humanitarian or refugee response plan coverage during the study period (see **Appendix 2**). |
| Outcomes | The primary outcome measure was population level vaccination coverage for any of the antigens listed above. Secondary outcome measures included vaccination delivery metrics such as drop-out rates, reported caseload for vaccine-preventable diseases; and resilience metrics (variable according to the study). |
| Languages | Arabic, English and French |

### Outcome measures

The primary outcomes of interest in this study were population level vaccination coverage for the vaccines outlined in the aims and objectives section above (defined according to the WHO schedule). We also considered secondary outcomes in two categories: [i] vaccination delivery outcomes as defined by included studies but including rates and reported caseloads for vaccine-preventable diseases (VPDs); access to routine immunization (typically defined as the proportion of eligible children within a given time period in receipt of a particular antigen dose); drop-out rates (for multiple dose regimens); and [ii] system resilience indicators where these were given e.g. the presence of systems for protecting financing for vaccination, composite measures such as facility readiness metrics, available vaccine stock, workforce numbers and reserve and so on (with variations from study to study according to design and setting).

### Identification of studies

Keyword-structured searches were performed in MEDLINE, EMBASE and Global Health (all via Ovid), CINAHL (via EBSCOHost), the Cochrane Collaboration and WHOLIS. These were accompanied by targeted searches for grey literature through the UNICEF, Global Polio Eradication Initiative (GPEI) and Technical Network for Strengthening Immunization Services (Technet – https://www.technet-21.org/en/) websites, because of the role these organisations and initiatives have played in producing and collating evidence and technical guidance to support routine immunisation delivery including in humanitarian contexts. All literature searches were performed between 3^rd^ and 9^th^ September 2021. A sample search strategy is provided in **Appendix 3**.

### Selection of studies

Studies were independently screened for inclusion on title and abstract, and then full texts reviewed by two members of the research team working independently, using the criteria outlined in Table 1. The first stage of screening (title and abstract) for articles identified through established search engines was performed using Rayyan QCRI, a free web application to support the conduct of systematic reviews by researchers working remotely (27). For results obtained from the UNICEF, GPEI and TechNet websites, initial screening was performed in MS Excel because it was not possible to download full reference details in an appropriate format for use in referencing software. Full text screening for all sources was conducted exclusively using MS Excel, and with reference to article PDFs. At each step, disagreements were resolved based on discussion between the two members of the screening pair.

### Data extraction, assessment of study quality, and data synthesis

Data were extracted in duplicate from each included study using a pre-developed extraction template in MS Excel. The extraction template was structured around the context-mechanism-outcome triumvirate emphasised in guidance on realist reviews (22,28). The template gathered basic study characteristics, general features of the intervention context (including the type of humanitarian crisis, and whether the intervention was geared towards prevention or outbreak response), the target disease; data on intervention structure using an approach informed by the WHO’s health system building blocks but including additional components such as service-user-focused aspects (demand management approaches such as community mobilisation and communications initiatives to bolster vaccination uptake); and finally measured outcomes. Importantly, the tool focused specifically on how inputs contributed to absorption, adaptation or even delivery system transformation (i.e. the intervention mechanism). Findings were synthesised using a thematic approach informed by the framework given in Figure 1.

Study quality was assessed independently using the Mixed Methods Appraisal Tool (MMAT), a validated tool developed which has been developed to help facilitate appraisal of public health research studies with multiple study designs and interventions within a single framework (29). Results from the duplicate extractions were inspected and areas of disagreement resolved by discussion between the two authors engaged in the extraction.

## RESULTS

A total of n=50 studies focused on childhood vaccination delivery were included after screening (see the flowchart in Figure 2, summary of results in **Appendix 4**, and quality appraisal summary in **Appendix 5** for further details). Included studies addressed interventions implemented in a range of settings, but the largest number of studies were from Nigeria (n=15, 30%), followed by South Sudan and Afghanistan (n=4, 8%), and Cameroon, Haiti and Somalia (n=3, 6%). Four studies (8%) addressed interventions applied across multiple settings. There was considerable diversity in health system and wider crisis contexts in which interventions were being implemented. Considering crisis settings, over half of the included studies described interventions in settings of active conflict or that were immediately post-conflict (n=27, 54%); a third were implemented in complex humanitarian emergencies including large-scale population displacement (n=18, 36%); and two studies (4%) focused on interventions implemented in the aftermath of natural disasters (Figure 3). Further detail on contextual settings is provided below.

**Figure 2.**
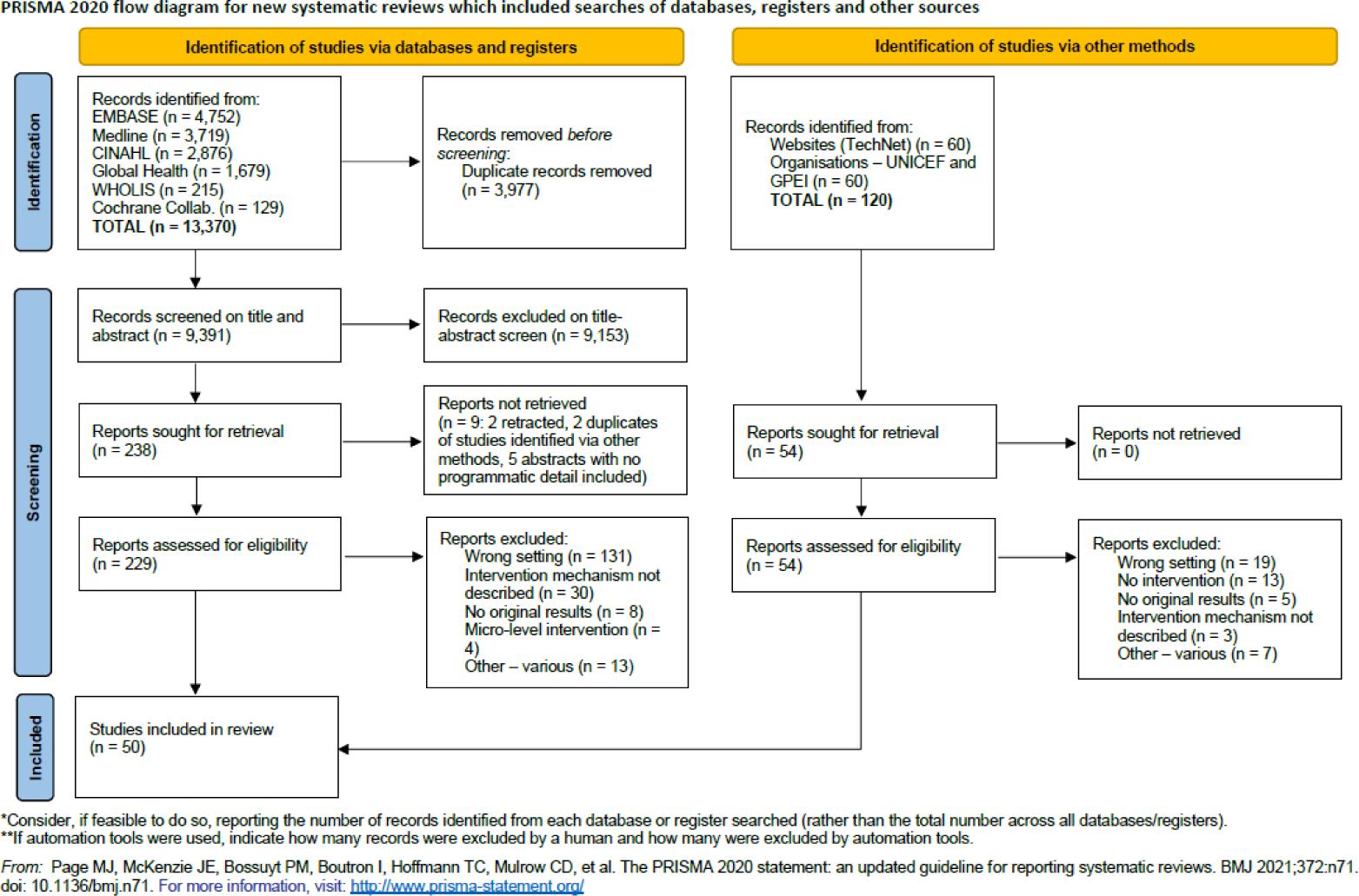
PRISMA flowchart describing the results of the screening and article selection process. In this review, the selection process is described in two flows; one relating to peer-reviewed literature sourced through formal databases (the left-hand stream), and one describing selection of peer-reviewed, grey and other sources identified through searches of organisational and other websites relevant to vaccination delivery (right-hand stream).

**Figure 3.**
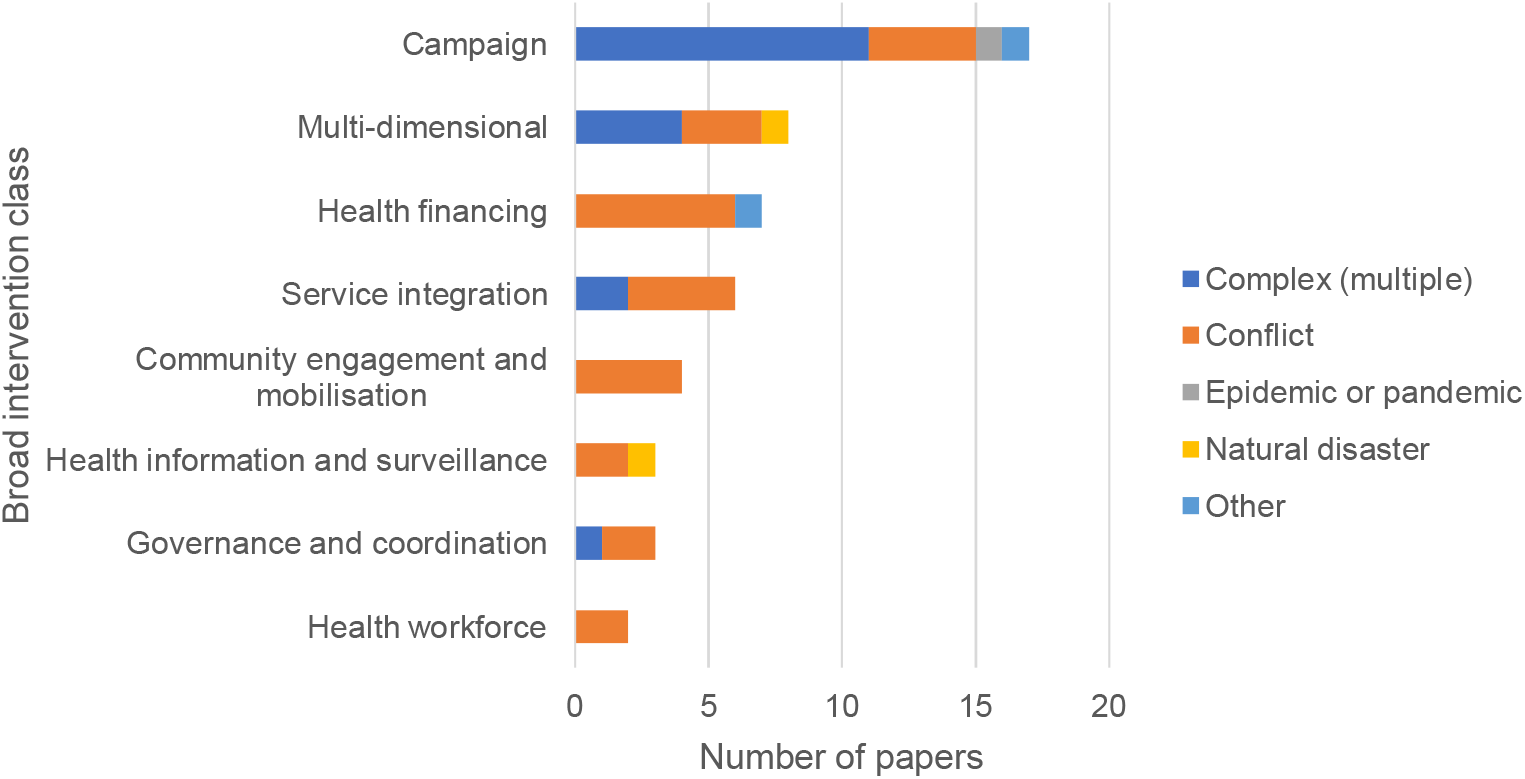
Breakdown of included articles by broad intervention class and the type of humanitarian context described. Campaigns were the most-commonly described interventions, and predominantly from complex humanitarian crisis settings.

We identified eight broad intervention classes (Figure 3), of which the commonly described were vaccination campaigns (n=17, 34%). Eight studies (16%) addressed multi-dimensional interventions, some of which included campaigns as one aspect of what was delivered but might also incorporate community mobilisation activities and governance or surveillance system strengthening among other activities. Health financing interventions were addressed in 7 studies (14%), including an evaluation of the effects of aid on immunization-related outcomes, and 2 studies on the application of national performance incentive policies implemented at the level of facilities or health workers. Community engagement activities were also considered in 4 studies (8%). Other papers addressed more narrowly focused interventions such as service integration (n=6, 12%), those geared towards improving governance coordination in vaccination delivery (n=3, 6%), health information (n=3, 6%) and health workforce (n=2, 4%). Finally, most studies addressed either interventions targeting multiple antigens from the routine schedule (n=17, 34%) or those aimed at improving coverage of different classes of poliomyelitis immunisation (n=19, 39%). A smaller number considered cholera or measles vaccination specifically.

A large majority of included studies (n=29, 58%) were mixed-methods program evaluations, but methodological approaches across the complete set were diverse. We did not identify any systematic reviews that explicitly addressed vaccination delivery in humanitarian settings and met the inclusion criteria for the study – specifically with regard to intervention design and delivery. This partly reflects a primary focus of this study on gathering information on intervention mechanisms, as well as measured effects. Significant methodological limitations were identified across almost all of the included studies (see summary MMAT judgements in **Appendix 5**).

Table 2 provides a summary of the principal intervention types identified in the review, the mechanisms by which these were seen to act (in all cases either adaptive, transformative or potentially transformative), a summary of relevant outcomes, and some of the key contextual modifiers in each case. A study-by-study summary of results is given in **Appendix 4**. As Table 2 shows, most included interventions supported system resilience through adaptation. Although reported outcomes varied according to the study, some important contextual modifiers were identified that cut across a number of studies, including: the level of insecurity in the operating environment; the level of population trust in providers; and the existence of pre-existing health system infrastructure (especially linked to polio eradication activities in security-compromised areas) around which to implement new interventions. The remainder of the results section is organised according to broad intervention class, beginning with vaccination campaigns. In each section, details of intervention mechanism are given alongside relevant contextual information to help explain how and why the intervention took the form that it did.

**Table 2.** Summary of interventions included in the review, the mechanisms by which these supported system resilience, outcomes and key contextual modifiers where these were identified. Multi-dimensional interventions are not included in this table because they comprise multiple discrete interventions acting in a variety of ways – please refer to the main text for further information on these.

| **Intervention class** | **Intervention type** | **Mechanism** | **Outcomes** | **Contextual modifiers** | **Relevant studies** |
| --- | --- | --- | --- | --- | --- |
| Campaign |  | Adaptive. | Variable according to campaign structure approaches (see appendix 4). All studies noted increases in coverage post-campaign but with variations according to geography or population group. | Wide variety of contexts described. Active conflict/insecurity strongly influenced choice of service delivery modality, in particular the extent to which community mobilisers were involved in demand-generation activities. | (30–46) |
| Health financing | Payment for performance | Adaptive | Variable evidence of effects on coverage for selected antigens depending on the study – one showing a positive effect, one showing no effect, and one showing a marginal decrease in coverage over time. | All included studies applied in conflict-affected settings. (Positive) wider health system financing environment likely to have influenced outcomes in one of these studies (50). | (50–52) |
|  | Funding disbursement | Adaptive | Rapid decline over time in difference between funds disbursed to health workers and funds accounted for. | Applied in conflict-affected setting only. Availability of a partner bank in-country with capacity to support mobile phone-based funding disbursement important to intervention delivery. | (53) |
|  | Private sector engagement | Transformative | Statistically significant increase in coverage for selected antigens in intervention area by comparison with controls. | Community-embeddedness of the intervention – through engagement of local councils – was important in supporting demand for vaccination through the public-private partnership. | (49) |
|  | Development financing | Adaptive | Improvements in coverage for selected antigens over time following uplift in macro-level financing. | Relevant countries included in both studies were primarily conflict-affected, with significant and ongoing disruption to health service delivery. | (47,48) |
| Service integration | Mobile Health Teams | Adaptive | Improvements in coverage for selected antigens over time but with variations across population groups. | Both interventions applied in conflict-affected settings. Existence of established governance mechanisms (linked to agreed basic package of care in one study, and a strategic plan and accountability framework in the other) | (54,55) |
|  | Nutrition and routine immunisation | Adaptive | Increases in number of children immunised in one study (coverage not reported) and coverage for selected antigens in the second study following implementation. | Both studies implemented in South Sudan in context of ongoing instability. Existing service integration policy identified as important success factor for the intervention in supporting pooling of funds across service domains. | (56,57) |
|  | Polio eradication and routine immunisation | Potentially transformative | Increases in coverage for selected antigens following implementation of the integrated interventions. | Both interventions implemented in conflict-affected contexts. Ability to mobilise sufficient financial resources to support delivery identified as a key success factor, as well as existence of established polio eradication architecture on which to capitalise. | (58,59) |
| Governance and coordination | Civil-military engagement | Potentially transformative | Improvements in accessibility for targeted areas noted following intervention implementation, as well as reductions in number of zero-dose children. | Both interventions applied in conflict-affected settings. Existence of governance mechanisms linked military leadership at regional level and the polio eradication programme in each country identified as an important factor in promoting civil-military engagement. | (60,61) |
|  | Cross-border coordination | Transformative | Improvements in vaccination coverage, case ascertainment for AFP among high risk populations identified following intervention implementation. | Political commitment from country governments and brokering by WHO identified as important to success of intervention. Contextual challenges tempering effects included ongoing population movement from South Sudan and local factors including a health worker strike in Kenya. | (62) |
| Health workforce | Volunteer community mobilisers | Adaptive | Reduction in the number of missed opportunities for polio vaccination following intervention introduction. | Supportive infrastructure of the polio eradication initiative in-country identified as key to success of the VCM intervention, as was population-level trust in these providers. | (63) |
|  | Technical surge capacity | Adaptive | Reduction in both missed opportunities for polio vaccination and documented polio cases following intervention implementation. | Substantial funding (in this case from BMGF) identified as an important contextual factor supporting the intervention, as well as prior implementation of a performance and accountability framework in WHO country offices that supported assessment of performance against surge personnel contracts. | (64) |
| Health information and surveillance | Monitoring and planning | Adaptive | Variable according to the intervention – see appendix 4. Reduction in the number of geographical locations with zero vaccination coverage by comparison with control areas in one (GIS-based) intervention. Effectiveness of rapid monitoring for campaign delivery unclear. | Prior experience of the polio eradication initiative with deployment of GIS-based population mapping supported intervention implementation. Continuing population movement undermined the effectiveness of rapid monitoring for campaign targeting in the second study. | (65,66) |
|  | Outreach surveillance | Adaptive | Reduction in reported polio case load, and improvements in performance against a series of AFP surveillance criteria. | Applied in a single post-conflict setting only. | (67) |
| Community engagement | Community mobilisation activities | Adaptive | Increases documented in vaccination coverage, or reductions in missed opportunities for vaccination depending on the study. | All four studies were implemented in northern Nigeria. An important factor influencing programme success in at least two of the studies included the long-established position of the polio eradication initiative and the supporting infrastructure it provided around which to build community mobilisation activities. Population trust in providers was also a key factor influencing success. | (68–71) |

### Supply-side interventions

#### Vaccination campaigns

A total of 17 studies in this category met the inclusion criteria, most (n=10) describing campaigns delivered in complex emergency settings, and a large proportion (n=7) focused on cholera prevention or control, with measles (n=4) and polio (n=3) also commonly targeted pathogens. Most studies were either post-hoc campaign evaluations (n=9) or cross-sectional surveys assessing campaign impact (n=8). These studies were distinguished from material on supplementary immunisation activities (SIAs) by the duration of engagement during the intervention, with an understanding that campaigns were short – up to a maximum of a few months in length for each individual round. Included articles described campaigns delivered in a broad range of country contexts, with the best-represented country again being Nigeria (n=4), and two studies from Cameroon.

All vaccination campaign studies described short-term and adaptive responses to crisis, including mobilisation of significant additional resources (financial, human and other) domestically and from international donors and other non-governmental actors. Those studies reporting the largest effects all concerned multi-component campaigns involving vaccination delivery through multiple service delivery modes (fixed site, mobile clinics and sometimes mass vaccination sites), but accompanied by community mobilisation activities, health worker recruitment and training, and support to cold chain improvement, among other interventions. For example, two linked studies addressing different aspects of the same cholera vaccination campaign among displaced persons in Borno, in northern Nigeria, showed 90% (95% CI 88-92%) first dose, and 73% complete (68-77%) OCV coverage in the target population following a multi-dimensional intervention involving door-to-door and fixed-site delivery modes, multiple information dissemination and communication routes (word-of-mouth, flyers, announcements and media spots) and health worker capacity building. However, the two studies also note important contextual factors contributing to success: the campaign benefited from a long-standing partnership governance model in which the Ministry of Health led but agencies and NGOs supported on the ground delivery, and preparedness in two key areas before the outbreak: training in cholera preparedness that had incidentally been run by the Nigeria CDC earlier the same year, and the licensing of OCV for use in Nigeria in the months prior to the outbreak (30,31).

These studies also emphasised the importance of prior networks built through polio eradication work in Northern Nigeria, and the ability of campaigns to capitalise upon this. Two further evaluation studies from Nigeria, considering polio and measles campaigns respectively, documented the use of polio eradication infrastructure to support short-term activities, including the use of existing GPEI governance structures, outreach capabilities especially in less secure areas that had been developed previously in partnership with the Nigerian military, and the integration of measles surveillance within case-based information gathering on acute flaccid paralysis (AFP). In these cases, near-term, adaptive responses built on long-term capacities in the vaccination delivery system in Nigeria intended to bolster resilience against the spread of poliomyelitis, although the results reported by the first study should be treated with caution because administrative coverage estimates exceed 100% (32,33).

Two studies from Cameroon considered campaign rollout for cholera and meningococcal vaccines against the backdrop of COVID-19 spread nationally. These studies emphasised the importance of central coordination through the Ministry of Health, the recruitment and training of cadres of vaccination workers (including community mobilisers to enhance outreach into remote and insecure communities) and the use of multiple communication modes with target populations – including through community leaders – to enhance uptake. Both studies reported large improvements in administrative vaccination coverage following the respective campaigns, but noted large regional variations. Neither study included a formal cross-sectional survey or time-series analysis to statistically evaluate campaign impact, so results should be treated with caution (34,35).

Drawing transferable lessons from other campaign-focused studies in this review is limited by the diversity in contexts in which they were implemented, ranging from multi-vaccine delivery in low-income settings in the midst of active conflict (36,37), to OCV and MMR deployment among displaced Rohingya refugees in Bangladesh (38). However, common strategies emerge across a number of these studies, including the use of multiple service delivery pathways (39–43), intensive community engagement and mobilisation activities (36,39,40,44,45), and central coordination led by the domestic Ministry of Health (36,38,40,41,46), which in combination were intended to increase vaccination coverage by mechanisms including bringing services closer to users, enhancing awareness of the need for vaccination and trust in service providers.

#### Health financing

Seven studies considered health financing interventions that ranged in focus from adaptive capacity building, to potentially transformative activities. These included two that looked at macro-level financing – specifically the value or otherwise of development aid in promoting improved health outcomes, including for vaccination, and system-strengthening in Gavi supported countries (47,48); one study that considered private sector provider engagement in routine immunisation provision (49); and four that considered in one way or another improvements to local level incentives for vaccination delivery (50–53). Of these four, three studies considered explicit incentivisation to facilities or health workers via pay for performance (P4P) (50–52); and one considered improvements to disbursement of programme funds to local level to promote vaccination delivery as a means of reducing the risk of delayed payment to HCWs (53). Most of these studies were from low-income settings in Sub-Saharan Africa, and 5 of the 6 considered interventions in conflict or post-conflict environments. The overall picture from these studies was mixed, as outlined below.

### Pay for performance (P4P) and financial disbursement mechanisms

Studies on P4P and disbursement mechanisms involved changes of a potentially transformative nature for system resilience by changing structures and accountability systems governing financing for vaccination delivery. Of the three studies that looked at P4P, two were from time-series analyses from Burundi (50,51) in the near-post conflict period, and one from Afghanistan (52) was a high-quality cluster RCT in the context of an ongoing, complex humanitarian crisis. All involved programmes overseen by the Ministry of Health in each country, but with contracting arrangements to facility level involving humanitarian actors and NGOs in service delivery. One study from Burundi found evidence of an improvement in completion rates for routine vaccination courses, especially for children from poorer households, after the introduction of the intervention (50), but the other two studies found no meaningful effect on vaccination uptake overall. The difference in measured effect may be partly down to intervention design: in Afghanistan incentives were provided directly to healthcare workers, whereas in Burundi payment was issued to facilities. Results in the Burundi study showing a positive effect on completion were also potentially influenced by context, because the intervention was implemented at the same time as a large overall increase in facility level budgets (for which the authors were unable to control) (50).

The study on disbursement mechanisms did not consider any of our primary outcomes, but did address intermediate effects (secondary review outcomes) including successful completion of health worker payments, widely understood to be a key operational challenge for programme delivery in LMICs in general, and humanitarian settings in particular (53). In this intervention, the WHO oversaw direct disbursement to the local level through a variety of activities including a newly introduced e-payment system, the establishment of close-to-facility payment sites and later the use of mobile payment systems, with reconciliation of funding distribution through partner meetings and other forms of information exchange. The intervention mechanism was two-fold: [i] to increase funding availability at the local level to support service delivery, and [ii] improve health worker satisfaction and motivation to deliver vaccination, as a result of prompt payment. In this observational study, the number of HCWs successfully paid increased markedly over the course of the programme, and match-up between funds disbursed and accounted for trended to 100%. This scheme was eventually extended beyond polio, to other routine immunisation workers, to help contribute to long-term system resilience.

### Private sector engagement

One study considered the integration of private providers into routine immunisation delivery through a public-private partnership (PPP) in Uruzgan province in central Afghanistan, which had historically suffered from both profound insecurity and chronic underinvestment in primary health care services (49). This cross-sectional analysis found a statistically significantly greater uptake of polio, DTP and measles vaccinations in intervention locations targeted by the PPP by comparison with control areas where access was primarily through mass vaccination campaigns. Involvement of private providers was promoted through a wide-ranging support package including payments to private practitioners (which did not appear to be performance-linked), provision of vaccine doses and consumables, HCW training and financial other forms of support to improve facilities. Private providers were also wrapped into broader community engagement through local councils, to improve awareness of the new service offer.

### Development financing and vaccination delivery

Finally, one ecological study looked at the effectiveness of development financing for health in South Sudan, linking macro-level donor and government investment in health to outcomes at national level through a population survey. This study concluded that despite considerable domestic instability, donor funding to support HSS had resulted in statistically significant improvements in measles (11.2% improvement in coverage, +/- 4.2%, p<0.001), DTP3 (13.1 +/- 3.6, p<0.001) and all-vaccination coverage (11.3, +/- 3.0, p<0.001) over 5 years from 2011-2015, albeit from low levels (48). These global findings covered marked variation at sub-national level, for which the study could offer not mechanism-based explanation as data on information on service delivery models and partnerships at this level were not gathered.

#### Service integration

Six studies considered system adaptation through service integration – four from sub-Saharan Africa and two from South Asia (Afghanistan and Pakistan). Two studies evaluated the impact of Mobile Health Teams (MHTs) offering integrated PHC including vaccination delivery in Afghanistan and Nigeria (54,55); two looked at integration between nutrition and routine immunisation services in South Sudan (56,57); and the final two studies considered integration of routine immunisation with polio outreach activities in Nigeria and Pakistan (58,59). All bar one were observational studies; the final study was a high-quality cluster RCT from Pakistan (59).

### Use of Mobile Health Teams

Two studies on the use of MHTs both showed statistically significant improvements in vaccination coverage, although the nature of these effects varied. A well-conducted, case-control study in Afghanistan found significant improvements only in uptake of first dose measles vaccination (54) by comparison with control areas (p=0.02), whereas in Nigeria, a document review-informed programme evaluation found improvements in all-vaccination coverage and course completion rates (from 19-55% over the duration of the intervention) in infants aged 12-23 months (55). The mechanisms by which MHTs worked had much in common across contexts, including an element of population targeting (especially in Nigeria where MHTs were deliberately targeted to settlements in the North of the country that were deemed high-risk for further AFP cases); intensive community engagement and outreach activities (integrated into the intervention through the inclusion of community mobilisers into the MHT in Nigeria, but parallel to it in Afghanistan) to promote awareness of, and trust in, services provided; a system of regular visits over time to build local population trust, rather than one-off interactions; and transparency regarding visit scheduling so that service users knew when teams would be in their locality.

### Integrating childhood immunisation and nutrition services

Two low-quality, observational studies considered the effect of integrating nutrition and immunisation services through single access points in South Sudan (56,57). Both showed improvements in uptake for routine immunisation following integration, but statistical analysis was rudimentary. In the first of these studies integration was into nutrition clinics in an IDP camp setting (56), whereas the second involved EPI integration into primary healthcare clinics (PHCCs) where this service had not previously been provided (57). The community-focus of these integration efforts was important: in the second study, pentavalent vaccination uptake was 23% greater (rate ratio 1.23, 95% CI 1.12-1.36) in PHCCs by comparison with paediatric outpatient departments (the control sites), suggesting a service-user preference for close-to-residence delivery settings.

### Integrating polio eradication and childhood immunisation services

Two studies addressed integration of routine immunisation and polio services. One of these, a high quality, cluster randomised trial of an integrating routine and polio immunisation activities, compared a control arm offering standard services, with two intervention arms offering an integrated programme of activities including community mobilisation and the use of additional fixed service delivery sites was used in security-compromised areas of Pakistan (the difference between intervention arms was chiefly in the polio vaccine formulation used – OPV in one arm, and IPV in the second). This study showed statistically significant improvements in the proportion of fully vaccinated children (7·3% [95% CI 4·5–10·0] increase in one arm vs control; and a 9·5% [6·9–12·0] increase in the second arm vs control). In both the intervention arms, the key mechanism changes were [i] the provision of multiple service delivery pathways (collaterals) and [ii] the use of intensive community mobilisation activities that were delivered in a culturally sensitive way (59). The second study, a programme evaluation from Nigeria considering the integration of routine immunisation with polio eradication work, also emphasised culturally-appropriate community outreach, through female community volunteer mobilisers, and saw a rise in the proportion of fully immunized children in the catchment areas from around 18% to 49% over the term of the intervention (58).

#### Governance and coordination

Three studies considered governance and coordination activities, two emphasising adaptive capacity through civil-military engagement, and the final study describing transformative capacity change through inter-governmental cooperation.

### Civil-military engagement for improved vaccination coverage in the context of insecurity

The two studies on civil-military engagement both addressed the use of military and/or security personnel to improve access and polio vaccination uptake in security compromised areas of Angola (60) and Nigeria (61) respectively, against a backdrop in both countries of enduring wild- and vaccine-derived polio circulation (including localised outbreaks). Both studies reported increases in uptake and reductions in the number of missed opportunities for vaccination (MOVs) in these settings although methodological limitations especially in the first of these studies limit the extent to which these gains can be ascribed to the interventions themselves. The mode of action of these interventions also differed: in Angola, military personnel were directly recruited to the programme, whereas in Nigeria, a cadre of civilians who had been supporting the military through community engagement in areas affected by the Boko Haram insurgency in the North were used to reach out to potentially eligible individuals. The key hallmark of both these interventions was a “feet on the ground” approach to improving uptake including in areas of significant insecurity, and the use of “hit-and-run” approaches to vaccine delivery with security cover during periods of unrest.

### Cross-border governance coordination for VPD prevention and control

A final study considered a cross-border governance and coordination initiative between the Kenyan, Somalian, Ethiopian and South Sudan governments to reduce the risk of polio transmission in the context of ongoing population movement. This was essentially a preparedness and planning intervention in which, with the support of WHO, representatives from respective country Ministries of Health met on a regular basis to identify important formal and informal population crossing points and transit hubs, agree on selection and recruitment of immunisation staff, and develop materials for workforce training and surveillance strengthening, among other activities. Identified improvements in vaccination coverage, strengthened cross-border situational awareness were reported (62). This could be described as a transformational change in resilience capacity at regional level by introducing and formalising new spaces for cross-border coordination and knowledge exchange that had not previously existed (i.e. wholly new system structures), underpinned by improvements in situational awareness, and connectivity between partner ministries, as well as stronger cross-border planning for future potential outbreaks.

#### Workforce development, flexibility and surge capacity

Two studies focused on the contributions of workforce interventions to improving vaccination outcomes, operating at different levels: one on the value-added of technical surge capacity to support immunisation delivery at local level, and the second on the contribution of community volunteers in mobilising service users to take up vaccination (although these were addressed indirectly by a number of other studies, covered elsewhere in the results section). Both studies were from Nigeria and both focused on polio vaccination delivery (63,64). The first study considered ability to bolster surge capacity using variable terms and other contractual changes for national or zonal staff, to enhance technical support to local areas during vaccination deployment. The use of surge technical capacity in this way was linked to progressive improvements in coverage, substantial declines in MOVs (a decrease in the number of localities with >10% of children missed by campaigns from 21% in 2012 to 3% in 2015) and improvements in intermediate indicators (e.g. the frequency with which micro-plans were updated) (64).

The second study looked at volunteer community mobilisers (VCMs) in Nigeria to address persistently low uptake in security compromised areas, vaccine scepticism, and distrust in government and officials. In this study, VCMs had an integrative role including supporting routine immunisation, but also WASH and other intervention promotion. These were financed through stipends and other (material) incentives but the main livelihood source is elsewhere. This intervention showed evidence of improvements in coverage but also a sharp decline in MOVs (from 4.5% in 2014 to 0.8% in 2018) which the study attributes to high levels of trust in VCMs because of their social position, cultural sensitivity of their approach, engagement modes including house-to-house visits, and their ability to update programme coordinators with critical soft information on pockets of low uptake (63).

#### Health information and surveillance

Three studies focused solely or primarily on situational awareness through health intelligence and surveillance activities – although as noted elsewhere in the results section, many of the interventions in other categories involved surveillance/health intelligence-focused components. All three studies were concerned with mechanisms for strengthening situational awareness in contexts featuring population movement and/or ongoing insecurity that could undermine the efficacy of more conventional approaches.

Two of the studies considered systems for strengthening awareness for campaign delivery and evaluation. One examined the use of geographical information system (GIS) technology to support microplanning processes for a measles campaign in Nigeria in 2017-18, focusing particularly on applications in conflict-affected northern states where tracking of mobile populations was much more challenging than in stable areas. Specifically, GIS mapping was used to geo-locate population centres to inform the positioning of fixed-sites for the campaign, and resulted in a reduction in the number of wards with zero vaccination coverage by comparison with states where standard population estimation approaches were used. The mechanisms by which this intervention were thought to have worked included [i] more accurate enumeration of target populations especially in the context of ongoing population movement, and [ii] a clearer view of ward boundaries than conventional (hand-drawn) approaches – both of which improved microplanning accuracy (65). The second study looked at the use of rapid monitoring approaches to gauge coverage and help improve campaign targeting in post-earthquake Haiti (66). In this study, a convenience sampling approach was used to assess household uptake at regular interviews; findings were used to better target mop-up vaccination activities. This approach helped strengthen resource management during the campaign but proved only partially effective as a monitoring technique because of continuing population movement. For this reason, the authors contended that it would be better suited to chronic rather than acute crisis situations.

A final, descriptive, study from Iraq considered the use of outreach surveillance activities linked to AFP case ascertainment for polio, to help assess uptake and effectiveness of immunisation in Iraq. This intervention involved use of AFP outreach workers to gather soft intelligence on RI delivery to pinpoint areas where microplanning for delivery needed to improve (alongside quantitative health data through immunisation delivery systems), and to help build awareness of services available among affected populations (67). The authors documented improvements in routine immunisation uptake and reductions in reported AFP case numbers through the outreach system.

### Demand-side interventions: community engagement and mobilisation

Although – as the summary table in **Appendix 4** makes clear – the vast majority of studies referenced community engagement as a trust-building measure to a greater or lesser degree, four studies explicitly focusing on this aspect met the inclusion criteria for the review, all of which concerned multi-dimensional community mobilisation activities (68–70), and one of which combined community mobilisation activities with direct observation of polio vaccination by officials to ensure proper administration of vaccination (71). All four were implemented in Nigeria, and most were concerned with improving polio vaccination uptake. Community mobilisation activities in Nigeria all took place in districts in the north and east of the country, against a backdrop of the Boko Haram insurgency, and all involved a strong element of cooperation with local community and religious leaders to build trust with affected communities. These complex interventions involved multiple outreach strategies including the use of roadshows, in-kind incentives (e.g. provision of soap or detergent, foodstuffs), health camps, and communication via multiple channels (69), agreement of mutually convenient vaccination point locations with community leaders (70) among other activities; a number incorporated recruitment and training of VCMs as part of the intervention (68). These strategies were used to encourage service use through demand-generation activities, reducing access barriers by positioning access points in areas where service users were more likely to take them up, and by enhancing trust in service providers.

Study outcomes focused predominantly on measures of administrative coverage and missed opportunities for vaccination. Three of the studies relied on narrative assessments of impact without formal statistical testing (68–70); the final study presented a rudimentary time series analysis (71). Two of the four studies found declines in the proportion of children who had not received any OPV doses during the study period (68,71); another showed concurrent, large increases in pentavalent vaccination coverage from an integrated intervention designed to improve uptake in hard-to-reach communities in Northern Nigeria (70), albeit with marked variations across geographies.

### Cross-cutting and multi-component interventions

Eight studies addressed sustained, multi-dimensional interventions. These interventions also tended to act at multiple levels, spanning macro-level governance and coordination changes, through to health worker training, and community outreach activities. Three of these studies looked at integrated, strategic interventions in response to poliomyelitis outbreaks, all in conflict-affected settings (72–74) and all featuring vaccination campaigns as part of the overall package. All emphasised the importance of strengthened coordination between key actors. A study from Somalia documented the introduction of a national polio control and coordination room to bring together key partners, improve communication and provide a framework for information/intelligence sharing; national coordination mechanisms were important in Ukraine and Syria (73,74); and regional mechanisms were used in Middle East and North African (MENA) countries for the 2017-18 polio outbreak response (75). Multi-modal social mobilisation activities were also central to achievement of improved outcomes.

Of these three studies, two considered regional activities in response to polio outbreaks in conflict-affected countries in the Middle East and North Africa, both involving multi-phase response plans not just to interrupt initial transmission, but then to focus on high-risk areas for importation. These evaluation reports indicate that through intensive community engagement it is possible to raise vaccine coverage even in areas of profound insecurity and constant population movement, but also that focused surveillance is really important in allocating resources appropriately and that multiple data points were triangulated for this (74). The first of these studies emphasised importance of targeting in identifying AFP hotspots, and the contribution of electronic syndromic surveillance systems in picking up emergent VPD case clusters and improving situational awareness (74). Results from the second study indicate that intermittent supplementary immunisation activities (SIAs) can be supportive of long-term system capacity by refreshing training and other essential functions and bolstering support for community outreach and surveillance initially built during an acute outbreak response, although these effects tail off over time (75). This study noted that gradual degradation of systems since the first SIA in 2013-14 may have contributed to the risk of re-emergence of polio cases in Syria in 2017-18 (75).

A further four studies looked at integrated, preventive responses, three focused on SIAs (76–78) and one on a district-level, NGO-led intervention to increase cholera vaccination uptake in urban slums in Haiti following the 2010 earthquake (79). SIAs achieved their measured effects through a combination of community mobilisation activities, capacity building of local health staff, mobilisation of funds from multiple sources (including international donor support) and supply chain strengthening activities. In Somalia, improvements linked to the SIAs contributed to long-term cold chain strengthening for all routine immunisation delivery (76). The district-level intervention in Haiti comprised multiple components including the introduction of new governance mechanisms (a coordinating committee), a communication plan, and re-allocation of the majority of NGO staff to supporting vaccination delivery for the duration of the intervention. This adaptive response to low coverage in their target communities benefited from an ability to surge staff from other areas – a disruptive approach for the NGO’s broader activities which is unlikely to have been sustainable beyond the term of the intervention (79).

## DISCUSSION

### Summary of key findings

This review identifies a series of interventions that may support vaccination system resilience capacities for improved routine childhood vaccination coverage (some of which also addressed catch-up vaccination delivery for older children) in settings of protracted humanitarian crisis. Most of these interventions reinforced adaptive resilience capacities but some (especially governance and workforce interventions) had transformative potential even if study results did not necessarily indicate radical change had been achieved within the term of the study. Although the diversity of settings described in this review precludes easy generalisation regarding important contextual factors, more successful interventions relied on leadership from domestic ministries of health, funding and – importantly – flexibility from agency and donor partners, and an ability to negotiate safe access for vaccinators and outreach workers.

Considering adaptive capacity, vaccination campaigns were the most extensively evidenced interventions and while their effects may appear to be short-lived, many of the examples considered here both built on existing capacities and contributed to development of new ones in a range of areas, including through introduction of novel governance structures and workforce capacities developed during previous activities. The most successful campaigns were multi-dimensional interventions that incorporated a mix of service delivery approaches (fixed-site, mobile team and mass-vaccination sites), intensive community mobilisation efforts, health worker training, and supply chain strengthening work. They were also often multi-phased to help both break chains of transmission (where the primary strategic objective was outbreak control) and prevent future outbreaks. The long-term sustainability of capacity strengthening through campaigns necessarily depends, however, on mobilisation of funding and other resources beyond the intervention lifecycle – some of which can be addressed by sustaining intervention through SIAs. None of the studies exclusively focused on campaigns offered detail on sustainability planning post-intervention.

Other adaptively-focused interventions included service delivery changes such as service integration and the use of MHTs. All included studies on service integration demonstrated improvements in vaccination coverage and course completion rates albeit with varying effect sizes and in studies of generally low quality. MHTs seemed to improve vaccination coverage by enabling outreach especially into poorer and more marginalised communities, by improving trust through regular interactions and the supporting activities community mobilisers drawn from the communities they served.

Some common mechanisms emerged across the higher-impact interventions identified in the review. For example, strengthening trust and increasing the range of access points were key themes especially among communities living in security-compromised zones, and many interventions directly addressed this through outreach models including the use of community volunteers, messaging through community (including religious) leaders, but also more controversially through cooperation with security personnel. Political and ethical challenges revolving around the deployment of security personnel in support of routine vaccination are considerable and this route is unlikely to be feasible or desirable in all settings. On the other hand, community volunteers were frequently identified as critical in tackling scepticism towards government and officials, but also contributed to bolstering situational awareness through improved case ascertainment for VPDs (as a linked, situational awareness-improving function).

Finally, a large proportion of the papers related directly or indirectly to lesson learning from polio eradication, and the resilience-building contributions PEI infrastructure could make with respect to routine immunization more generally. Particular areas of learning included the use of multi-modal approaches to vaccination and surveillance in security-compromised areas, the value and use of existing outreach networks into communities, and messaging strategies. Objectives for polio control (specifically eradication) are very different to those for many other VPDs where primary aims are more likely to be interruption chains of transmission in the context of outbreaks or to keep the burden of mortality and morbidity low in the face of endemicity (for diseases such as measles for example). This should not, however, preclude the use of infrastructure developed for polio control to support wider routine immunisation objectives.

Evidence in some areas was notable by its scarcity. For example, material on governance and financing was both limited and showed conflicting evidence on vaccination-related outcomes, although some promising interventions were identified in these domains, including the use of direct disbursement mechanisms for health worker payment, and a model of cross-border cooperation in the Horn of Africa to support preparedness and planning to reduce poliomyelitis risk. In addition, while some studies touched on leadership and oversight capacity development, we identified no studies that explicitly focused on leadership models contributing to resilience capacities for routine immunisation. Similarly, evidence on governance reforms such as decentralisation, which have been implemented in some eligible countries (e.g. Kenya and Tanzania) was not forthcoming, although one study briefly considered the importance of decentralised decision-making as a contextual contributor to the success of a nutrition-immunisation integration intervention (57). Finally, although we did not specifically screen to include studies on cost and cost-effectiveness, data on costs associated with intervention implementation were conspicuously absent from included studies. This is a notable shortcoming given the fundamental importance of sustainable financing for health system resilience especially in crisis-affected settings.

A number of systematic reviews have been published in recent years addressing the effectiveness of interventions in humanitarian settings, some including data on vaccination delivery (80,81), and there is now a large body of evidence considering the effectiveness of interventions to improve vaccination coverage from community level upwards in low- and middle-income settings more generally (e.g. 16). This review is, however, distinctive in its macro- and meso-level interventional focus, in employing a realist perspective to understand how, where and why interventions may be effective, and its emphasis on gathering data from protracted crisis settings. Many existing studies and guidance documents focus on acute-phase responses without consideration to ways in which these may support, or undermine, long-term capacity within the system to respond to changing circumstances on the ground, or the mechanisms by which they may do so (17,83,84).

Nevertheless, many of the central messages from this review – including the value of recruiting local staff to improve trust and vaccination uptake, flexibility in service delivery modes (including the use of mobile services), and use of electronic systems to strengthen supply chain management and health intelligence – support findings from reviews and guidance elsewhere (81,83,85). Findings also suggest that SIAs, as integrative interventions, can have important and wide-ranging effects not just on vaccination uptake, but also on wider system capacity through e.g. workforce training, introduction of new governance and coordination mechanisms, and support to outreach activities. These effects should be balanced against potential undermining effects on routine immunisation through fixed centres that have been observed in some settings (86), but SIAs are likely to remain an important part of the adaptive armoury to expand coverage in humanitarian settings given their effectiveness at picking up children missed through routine delivery (87). In addition, a number of promising practices are identified, including the use of flexible contracting for healthcare workers and novel financial disbursement mechanisms, for which evidence remains limited but which nevertheless could address delivery problems widely acknowledged in these settings.

### Limitations of the review

Limitations to the findings reported here relate to both the nature of the underlying evidence base, and the way in which the review was conducted. As the quality appraisal results show (see **Appendix 5**), most included studies were observational works with significant methodological limitations. This particularly affected descriptive, quantitative analyses included in the review (most of them program evaluations). Outcome reporting was in general poor: measures were incompletely described and frequently related weakly to study objectives or included no clear baseline data against which to measure effects. The quality and detail of intervention description was highly variable, making it difficult to tell which particular intervention components were driving reported results.

Although we were careful to use explicit definitions to guide the review, clarity in the wider literature on some of the key terms is lacking and may have contributed to relevant results being missed. For example, we used a response plan-driven definition of protracted crises that assumes that the introduction of a RP coincided fairly precisely with the duration of the crises of interest. This is often not the case: there may be delays of up to several years before RPs are formulated and agreed – as in the case of the Syria Crisis response, for example. Secondly, an important goal of this review was to capture data from a wider range of sources than the peer-reviewed literature alone, to better capture emergent best practice. Structured searching of grey literature sources remains challenging and it is likely that some relevant material was missed.

As with all systematic reviews, our findings also cannot account for unpublished or negative results. This may explain the lack of data relating to absorption as a resilience mechanism – that they were not reported simply because they involved services performing in much the same way but more intensively (e.g. through changes to service opening hours, workforce redeployment etc). Because of the diversity of intervention types, study designs and study contexts included, it was neither possible nor desirable to produce summary statistics of intervention effects beyond those reported in the results section.

Finally, we noted imbalances in the geographical representativeness of studies included. On one hand, the balance of included evidence was skewed strongly towards unstable, low income settings. This is unsurprising given that most countries in protracted crisis today fall into this category, but it reduces the potential transferability to middle income countries hosting large displaced populations today. Evidence from countries such as Jordan, Iran and Turkey, for example – all of which host large populations displaced by conflict – was notably absent in this review. On the other, a large proportion of included studies were conducted in Nigeria – reflecting historical challenges with vaccine delivery and particularly polio control in the north of the country.

### Policy implications

Findings from this review suggest that no “silver bullet” solution exists to promoting resilience in vaccination delivery systems in protracted humanitarian crises, and adaptations are likely to be needed across a range of fronts to address significant access barriers (many of which stem directly from population displacement and chronic insecurity), low trust in service providers, and limitations to effective VPD prevention and control imposed by national borders. Periodic intensification of vaccination delivery via campaigns and SIAs is likely to be a mainstay of adaptive responses to crises whatever the context, to account for shortfalls in routine delivery. However, there is a strong steer from this work for recruitment of non-traditional workforce cadres from within affected communities – including community mobilisers – to help enhance uptake over the long-term, especially in security compromised areas where trust in government and agency representatives may be low.

Resilience in vaccination coverage is also likely to be enhanced through the concurrent use of multiple service channels to reach affected populations, including MHTs and integration with in-demand services such as nutritional support. However, there will inevitably be trade-offs in cost terms to expanded service availability in this way given resource constraints in many humanitarian settings, and the success of any of these interventions will ultimately depend on the willingness of domestic and international actors (including donors) to ensure stability in funding flows to crisis-affected countries.

## Conclusion

Strengthening the resilience of vaccination delivery systems in protracted humanitarian crises depends on system adaptation across a range of areas, including bolstering access through strengthened outreach, multiple service pathways and better integration with other essential services, as well as demand-generation activities. Future work should consider evidence not just on adaptive and transformative measures to support improvements in vaccination coverage in these settings, but also economic analyses given the significant resource constraints under which decision-makers in humanitarian contexts have to operate.

## Data Availability

All data generated or analysed during this study are included in this published article and its supplementary information files.

## DECLARATIONS

### Ethics approval and consent to participate

This review was based entirely on data from sources already in the public domain. No ethical approval was required.

### Consent for publication

Not applicable.

### Authors’ contributions

CRediT author statement:

SAI – conceptualisation, methodology, investigation, writing – original draft, writing – review and editing, visualisation, project administration, funding acquisition

LST – methodology, investigation, writing – review and editing

SB – conceptualisation, methodology, supervision, writing – review and editing FMF – conceptualisation, writing – review and editing

KB – conceptualisation, supervision, writing – review and editing JB – conceptualisation, supervision, writing – review and editing

