## Supplementary materials 1 - appendices 1,2,3,5,6 for "Strengthening vaccination delivery system resilience in the context of protracted humanitarian crisis: a realist-informed systematic review"

### Appendix 1: developing a working analytical framework for the review

Understanding of which interventions may bolster system resilience in health, and how remains limited partly because of enduring analytical differences over the way in which resilience is conceptualised (1,2). There is, however, a degree of consensus in the health literature on the kinds of *attributes* associated with system resilience, if not necessarily the means by which these can be enhanced. They include “hard” attributes such as availability of material and human resources, and the existence of collateral pathways (i.e. the existence of multiple mechanisms through which, for example, medical products or health services can be delivered) (3,4). Effective information management is also vital although the balance between formal surveillance and softer, more immediate data from human intelligence systems in shaping system responses has emerged as an area of debate in humanitarian settings and in the context of COVID-19 responses (5,6). “Soft” attributes include features such as networking and connectivity (7). Empirical studies of resilience governance and system leadership have been few including in health, although existing health research does identify attributes including legitimacy and knowledge management as important (8–10). These attributes may derive in part from the health system and wider context in which interventions operate, but also form part of activities linked to the intervention itself.

While the ultimate goal of vaccination delivery systems can be considered to be generating improvements in vaccination coverage, and thereby population health outcomes (including mortality and morbidity from VPDs), the attributes described imply a focus on interventions that act through intermediate pathways, such as improvements in the quality of health information, integration with other services and service providers, the scale and durability of resource inputs (human, financial and others) and so on. These, and other similar outcomes, form an important focus for this review: our focus was not just on vaccination delivery endpoints (population coverage, deaths and hospitalisations averted, for example) but also on the system outputs and outcomes through which these effects were achieved.

We therefore developed a working framework to enable a layered reading of included articles, exposing:

- Context: including both the broader national context and humanitarian crisis within which the intervention implementation occurred, as well as the background state of the health system (understood with reference to the WHO health system building blocks)
- Mechanism: as a function of intervention activities on the health system building blocks, the ways in which these supported the development of recognised resilience attributes (such as collateral service pathways, strengthened networking and other features highlighted above) and ultimately the specific mechanism (whether absorptive, adaptive, or transformative) by which resilience was strengthened as a result of the intervention.
- Outcomes: vaccination-related outcomes as set out in the methods section, and as reported by included articles.

The framework outlines a loose progression from inputs to outcomes (e.g. increased vaccination coverage) but does not make explicit assertions about causal links between individual elements, about feedback loops that are, in reality, likely to apply, or about the dynamic nature of the relationship between interventions and wider context. This is because the nature of these relationships is likely to vary from intervention to intervention.

#### *Appendix 1 references*

1. Turenne CP, Gautier L, Degroote S, Guillard E, Chabrol F, Ridde V. Conceptual analysis of

health systems resilience: A scoping review. Soc Sci Med. 2019;232:168–80.

### Appendix 2: list of eligible settings and their eligibility periods

Using data from UN OCHA, and the humanitarian information gathering portals ReliefWeb and Humanitarianresponse.info, the following countries were categorised as eligible for inclusion as protracted crisis settings during the study period. Note that not all listed countries were eligible for the entire study period; some were eligible only for specific time points (coinciding with periods when they were covered by a HRP or equivalent) – but that all countries had to have had a run of at least 5 years under a response plan of one kind or another to meet the criteria for a protracted crisis setting.

| ***Country*** | ***Eligible period(s)*** | ***Relevant response plans*** |
|---|---|---|
| *Core countries (with country-specific RPs +/- relevant regional RPs of which they formed part)* | | |
| Afghanistan | Full duration of the study | Consolidated inter-agency appeals since 1995-96 (and prior to this); consecutive HRPs from 2009-2021 |
| Angola | 2001-2002 | Consolidated inter-agency appeals, 1995-2002; DRC RRP |
| Burkina Faso | 2013-2021 | Consecutive HRPs from 2013-2021 |
| Burundi | 2016-2021 | Consecutive HRPs from 2016-2021; DRC RRP |
| Cameroon | 2014-2021 | Consecutive HRPs from 2014-2021 |
| Chad | 2004-2021 | Consecutive HRPs from 2004-2021 |
| Central African Republic | 2003-2021 | Consecutive HRPs from 2003-2021 |
| Democratic Republic of Congo | Full duration of the study | Consecutive HRPs from 1999-2021 |
| Ethiopia | 2017-2021 | Consecutive HRPs from 2017-2021 |
| Haiti | 2010-2021 | Consecutive HRPs from 2010-2021 |
| Honduras | 2014; 2021-22 | Drought emergency response 2014; HRP from 2021-22 |
| Iraq | Full duration of the study | Consolidated inter-agency appeals since at least 1995/6; consecutive Syria 3RPs 2015-2021; consecutive HRPs from 2014-2021 |
| Lebanon | 2015-2021 | Lebanon war response 2006-7; Syria 3RP 2015-2021; ERP 2021-present |
| Libya | 2011-2021 | Consecutive HRPs 2015-2021 |
| Mali | 2012-2021 | Consecutive HRPs from 2012-2021 |
| Myanmar | 2013-2021 | Consecutive HRPs from 2013-2021 |
| Niger | 2011-2021 | Consecutive HRPs from 2011-2021 |
| Nigeria | 2014-2021 | Consecutive HRPs from 2014-2021 |
| Occupied Palestinian Territories | Full duration of the study | UNRWA presence since 1949; Consecutive HRPs from 2003-2021 |
| Somalia | Full duration of the study | Consecutive HRPs from 1998-2021 |
| South Sudan | 2011-2021 (2010 if including HRP prior to independence) | Independence in 2011; Consecutive HRPs from 2010-2021 |
| Sudan | Full duration of the study | Consecutive HRPs from 1993-2021 |
| Syria | 2012-2021 | Syria HARP 2012 and subsequent domestic plans; 3RP from 2015 onwards. |
| Ukraine | 2014-2021 | Consecutive HRPs from 2014-2021 |
| Yemen | 2008-2021 | Consecutive HRPs from 2008-2021 |
| *Countries included as partners in regional refugee response plans only* | | |
| Bangladesh | 2017-2021 | Rohingya RRP 2017-21 |
| Congo | 2018-2021 | DRC RRP 2018-21 |
| Egypt | 2015-2021 | Consecutive Syria 3RPs 2015-present |
| Iran | Full duration of the study | Afghanistan RRP |
| Jordan | 2015-2021 | Consecutive Syria 3RPs 2015-present |

| | | |
|---|---|---|
| Kenya | 2008-2021 | Refugee response plan 2014-2020; Emergency HRP 2008-2013; South Sudan RRP |
| Pakistan | Full duration of the study | Afghanistan RRP |
| Rwanda | 2018-2021 | DRC RRP 2018-21 |
| Tajikistan | Full duration of the study | Afghanistan RRP |
| Tanzania | 2018-2021 | DRC RRP 2018-21 |
| Turkey | 2015-2021 | Consecutive Syria 3RPs 2015-present |
| Turkmenistan | Full duration of the study | Afghanistan RRP |
| Uganda | 2018-2021 | DRC RRP 2018-21; South Sudan RRP |
| Uzbekistan | Full duration of the study | Afghanistan RRP |
| Zambia | 2018-2021 | DRC RRP 2018-21 |

Abbreviations: 3RP (Syria Crisis only) = Regional Refugee and Resilience Plan; ERP = emergency response plan; HARP = humanitarian assistance response plan; HRP = humanitarian response plan; RRP = regional response plan for refugees.

### Appendix 3: sample search strategy

Sample search structure as applied in MEDLINE:

Ovid MEDLINE(R) ALL <1946 to September 03, 2021>

| | | |
|---|---|---|
| 1 | exp Immunization, Passive/ or exp Immunization Schedule/ or exp Immunization/ or exp Immunization, Secondary/ or exp Immunization Programs/ | 192796 |
| 2 | exp Vaccination/ or exp Mass Vaccination/ | 92277 |
| 3 | Vaccines/ or bacterial vaccines/ or toxoids/ or viral vaccines/ or cholera vaccines/ or diphtheria-tetanus vaccine/ or Diphtheria-Tetanus-acellular Pertussis Vaccines/ or Diphtheria-Tetanus-Pertussis Vaccine/ or Diphtheria-Tetanus Vaccine/ or Pertussis vaccine/ or Haemophilus Vaccines/ or Meningococcal Vaccines/ or BCG vaccine/ or tuberculosis vaccines/ or Heptavalent Pneumococcal Conjugate Vaccine/ or Pneumococcal vaccines/ or Papillomavirus vaccines/ or Human Papillomavirus Recombinant Vaccine Quadrivalent, Types 6, 11, 16, 18/ or Measles-Mumps-Rubella Vaccine/ or Measles Vaccine/ or Mumps Vaccine/ or Rubella Vaccine/ or Poliovirus vaccines/ or Poliovirus vaccine, inactivated/ or Poliovirus vaccine, oral/ or Viral Hepatitis Vaccines/ or hepatitis A vaccines/ or hepatitis b vaccines/ or Rotavirus vaccines/ or chickenpox vaccine/ or Influenza Vaccines/ or Typhoid-Paratyphoid Vaccines/ or Tetanus Toxoid/ or Yellow Fever Vaccine/ | 182764 |
| 4 | or/1-3 | 301617 |
| 5 | exp child/ or exp infant/ or mothers/ or women/ or pregnant women/ or female/ | 10199346 |
| 6 | 4 and 5 | 133861 |
| 7 | ((vaccinat* or revaccinat* or immunization or immunisation) adj3 (child* or infant? or newborn? or neonat* or baby or babies or toddler? or woman or women or mother?)).ti,ab. | 22878 |
| 8 | ((immunization or immunisation or vaccination) adj (rate* or coverage or uptake or adher* or complian* or drop-out or drop out or access*)).ti,ab. | 14272 |
| 9 | (((immunization or immunisation or vaccination) and (system* or service* or delivery or pathway)) adj (readiness or prepared* or responsive* or quality or safe* or resilien* or robust* or adapt* or absorb* or absorp* or transform*)).ti,ab. | 101 |
| 10 | 6 or 7 or 8 or 9 | 144976 |
| 11 | relief work/ or exp "warfare and armed conflicts"/ or refugees/ or exp disasters/ or genocide/ or ethnic cleansing/ or exp disasters/ or exp disease outbreaks/ or earthquakes/ or volcanic eruptions/ or floods/ or landslides/ or tidal waves/ or tsunamis/ or cyclonic storms/ or droughts/ or starvation/ or famine/ or disaster medicine/ | 311247 |
| 12 | (humanitarian* or protracted crisis or protracted crises or complex emergenc* or conflict-affected or conflict affected or fragile countr* or "fragile state* fragile and conflict affected" or "fragile and conflict-affected" or FCAS or insecur* or secur* or transition* countr* or internal displace* or internally displaced person* or displaced population* or mobile population* or forced migrat* or forced migrant or typhoon* or cyclone* or hurricane* or aid work* or financial crisis or economic crisis).hw,kf,ti,ab,cp. | 137200 |
| 13 | 11 or 12 | 436977 |
| 14 | Developing Countries.sh,kf. | 89235 |
| 15 | (Africa or Asia* or Caribbean or West Indies or South America or Latin America or Central America or Eastern Mediterranean or Americas or Western Pacific).hw,kf,ti,ab,cp. | 473656 |
| 16 | (Afghanistan or Bangladesh or Burkina Faso or Burundi or Central African Republic or CAR or Chad or Colombia or Democratic Republic of Congo or DRC or Eritrea or Ethiopia or Iraq or Lebanon or Mali or Myanmar or Niger or Nigeria or Pakistan or Peru or Somalia or Sudan or South Sudan or Syria or Syrian Arab Republic or Turkey or Uganda or Ukraine or Venezuela or Yemen).ti,ab. | 253763 |
| 17 | (Albania or Algeria or Angola or Argentina or Armenia or Azerbaijan or BBelarus or Belize or Benin or Bhutan or Bolivia or Botswana or Brazil or Bulgaria or Cambodia or Cameroon or Chile or China or Congo or Costa Rica or Cote dIvoire or Ivory Coast or Cuba or Democratic Peoples Republic of Korea or DPRK or North Korea or Djibouti or Dominica or Dominican Republic or Ecuador or Egypt or El Salvador or Gambia or Georgia or Ghana or Guatemala or Guinea or Guinea Bissau or Haiti or Honduras or India or Indonesia or Iran or Jamaica or Jordan or Kazakhstan or Kyrgyzstan or Kyrgyz Republic or Kenya or Laos or Lao PDR or Lao or Liberia or Libya or Madagascar or Malawi or Malaysia or Maldives or Mauritania or Mexico or Mauritania or Moldova or Mongolia or Montenegro or Morocco or Mozambique or Namibia or Nepal or Nicaragua or Republic of North Macedonia or North Macedonia or Palestine or Occupied Territories or Occupied Palestinian Territories or Gaza or West Bank or Papua New Guinea or Paraguay or | 962412 |

| | | |
|---|---|---|
| | Philippines or Russia or Russian Federation or Rwanda or Senegal or Serbia or Sierra Leone or Sri Lanka or South Africa or Tajikistan or Tanzania or Timor Leste or Togo or Thailand or Turkmenistan or Uzbekistan or Vietnam or Western Sahara or Zambia or Zimbabwe).ti,ab. | |
| 18 | ((developing or less* developed or under developed or underdeveloped or middle income or low* income or underserved or under served or deprived or poor*) adj (countr* or nation? or population? or world)).ti,ab. | 113426 |
| 19 | ((developing or less* developed or under developed or underdeveloped or middle income or low* income) adj (economy or economies)).ti,ab. | 697 |
| 20 | (low* adj (gdp or gnp or gross domestic or gross national)).ti,ab. | 292 |
| 21 | (low adj3 middle adj3 countr*).ti,ab. | 21684 |
| 22 | (lmic or lmics or third world or lami countr*).ti,ab. | 9521 |
| 23 | 14 or 15 or 17 or 18 or 19 or 20 or 21 or 22 | 1386105 |
| 24 | 13 and 23 | 63847 |
| 25 | (10 and 24) or (10 and 16) | 6413 |
| 26 | exp health policy/ or exp health services administration/ or health services/ or child care/ or personal health services/ or community health services/ or community health centers/ or health facilities/ or ambulatory care/ or universal health care/ or child health services/ or maternal-child health services/ or telemedicine/ or preventive health services/ or "health services needs and demand"/ or workforce/ or health workforce/ or community health workers/ or exp medical informatics/ or public health informatics/ or exp population surveillance/ or behavioral risk factor surveillance system/ or epidemiological monitoring/ or exp "equipment and supplies"/ or "equipment and supplies utilization"/ or leadership/ or health education/ or healthcare financing/ or financing, government/ or financing, personal/ or induced demand/ or vaccination promotion/ or vaccination campaign/ or communication/ or exp "Health Care Quality, Access, and Evaluation"/ | 9868768 |
| 27 | (strengthening or system strengthening or health system strengthening).ti,ab. | 35858 |
| 28 | (social mobili*ation or community mobili*ation or community outreach or community engage* or community health volunteer or outreach or mobile unit or mobile medical unit or mobile team or cash transfer or cct or cash incentive or demand generat*).ti,ab. | 28371 |
| 29 | (logistic* or deliver* or supply chain* or supply-chain* or cold chain* or cold-chain*).ti,ab. | 1073065 |
| 30 | (governance or accountab* or oversight or regulat* or stewardship).ti,ab. | 2053531 |
| 31 | 26 or 27 or 28 or 29 or 30 | 12104943 |
| 32 | 25 and 31 | 4604 |
| 33 | limit 32 to (yr="2001 -Current" and (arabic or english or french)) | 3719 |

### Appendix 4: summary table of included studies

**See separate MS Excel spreadsheet for Appendix 4 content.**

Notes:

AFP = Acute Flaccid Paralysis

BCG = Bacillus Calmette–Guérin vaccine (against tuberculosis)

BMGF = Bill and Melinda Gates Foundation

CHD = Child Health Day

CJTF = Civilian Joint Task Force (Nigeria)

DTP = Diphtheria, Tetanus and Pertussis vaccine

EPI = Expanded Program on Immunisation

EWARN = Early Warning, Alert and Response Network (surveillance)

GIS = Geographical Information System

GPEI = Global Polio Eradication Programme

HCW = Health Care Worker

ICG = International Coordination Group on Vaccine Provision

IPV = Inactivated Polio Vaccine

MCV = Measles-Containing Vaccine

MHT = Mobile Health Team

MMR = Measles, Mumps and Rubella vaccine

OCV = Oral Cholera Vaccine

OPV = Oral Polio Vaccine

OTP = Outpatient Therapeutic Programme (nutrition)

P4P = Pay for Performance

PBF = Performance-Based Financing

PCV = Pneumococcal Conjugate Vaccine

RI = Routine Immunisation

SIA = Supplementary Immunisation Activity

UNICEF = United Nations International Children’s Emergency Fund

VCM = Volunteer Community Mobiliser

WASH = Water, Sanitation and Hygiene

WHO = World Health Organisation

### Appendix 5: overall summary of quality appraisal results

| | | Yes | | No | | Can't tell | |
|---|---|---|---|---|---|---|---|
| | | n | % | n | % | n | % |
| **SCREENING QUESTIONS** | S1. Are there clear research questions? | 50 | 100% | 0 | 0% | 0 | 0% |
| | S2. Do the collected data allow to address the research questions? | 48 | 96% | 1 | 2% | 1 | 2% |
| **1. QUALITATIVE STUDIES (n=10)** | 1.1. Is the qualitative approach appropriate to answer the research question? | 10 | 100% | 0 | 0% | 0 | 0% |
| | 1.2. Are the qualitative data collection methods adequate to address the research question? | 8 | 80% | 0 | 0% | 2 | 20% |
| | 1.3. Are the findings adequately derived from the data? | 9 | 90% | 0 | 0% | 1 | 10% |
| | 1.4. Is the interpretation of results sufficiently substantiated by data? | 9 | 90% | 1 | 10% | 0 | 0% |
| | 1.5. Is there coherence between qualitative data sources, collection, analysis and interpretation? | 9 | 90% | 0 | 0% | 1 | 10% |
| **2. RANDOMIZED CONTROLLED TRIALS (n=2)** | 2.1. Is randomization appropriately performed? | 2 | 100% | 0 | 0% | 0 | 0% |
| | 2.2. Are the groups comparable at baseline? | 2 | 100% | 0 | 0% | 0 | 0% |
| | 2.3. Are there complete outcome data? | 2 | 100% | 0 | 0% | 0 | 0% |
| | 2.4. Are outcome assessors blinded to the intervention provided? | 1 | 50% | 1 | 50% | 0 | 0% |
| | 2.5 Did the participants adhere to the assigned intervention? | 2 | 100% | 0 | 0% | 0 | 0% |
| **3. NON-RANDOMIZED STUDIES (n=5)** | 3.1. Are the participants representative of the target population? | 4 | 80% | 0 | 0% | 1 | 20% |
| | 3.2. Are measurements appropriate regarding both the outcome and intervention (or exposure)? | 5 | 100% | 0 | 0% | 0 | 0% |
| | 3.3. Are there complete outcome data? | 4 | 80% | 0 | 0% | 1 | 20% |
| | 3.4. Are the confounders accounted for in the design and analysis? | 1 | 20% | 4 | 80% | 0 | 0% |
| | 3.5. During the study period, is the intervention administered (or exposure occurred) as intended? | 4 | 80% | 0 | 0% | 1 | 20% |
| **4. QUANTITATIVE DESCRIPTIVE STUDIES (n=35)** | 4.1. Is the sampling strategy relevant to address the research question? | 34 | 97% | 0 | 0% | 1 | 3% |
| | 4.2. Is the sample representative of the target population? | 23 | 66% | 0 | 0% | 12 | 34% |
| | 4.3. Are the measurements appropriate? | 33 | 94% | 2 | 6% | 0 | 0% |
| | 4.4. Is the risk of nonresponse bias low? | 13 | 37% | 5 | 14% | 17 | 49% |
| | 4.5. Is the statistical analysis appropriate to answer the research question? | 31 | 89% | 2 | 6% | 2 | 6% |
| **5. MIXED METHODS STUDIES (n=2)** | 5.1. Is there an adequate rationale for using a mixed methods design to address the research question? | 2 | 100% | 0 | 0% | 0 | 0% |
| | 5.2. Are the different components of the study effectively integrated to answer the research question? | 1 | 50% | 1 | 50% | 0 | 0% |
| | 5.3. Are the outputs of the integration of qualitative and quantitative components adequately interpreted? | 1 | 50% | 1 | 50% | 0 | 0% |
| | 5.4. Are divergences and inconsistencies between quantitative and qualitative results adequately addressed? | 1 | 50% | 0 | 0% | 1 | 50% |
| | 5.5. Do the different components of the study adhere to the quality criteria of each tradition of the methods involved? | 1 | 50% | 0 | 0% | 0 | 0% |

### Appendix 6: PRISMA checklist

| Section and Topic | Item # | Checklist item | Location where item is reported |
|---|---|---|---|
| **TITLE** | | | |
| Title | 1 | Identify the report as a systematic review. | 1 |
| **ABSTRACT** | | | |
| Abstract | 2 | See the PRISMA 2020 for Abstracts checklist. | 2 |
| **INTRODUCTION** | | | |
| Rationale | 3 | Describe the rationale for the review in the context of existing knowledge. | 3 |
| Objectives | 4 | Provide an explicit statement of the objective(s) or question(s) the review addresses. | 3 |
| **METHODS** | | | |
| Eligibility criteria | 5 | Specify the inclusion and exclusion criteria for the review and how studies were grouped for the syntheses. | 4, Table 1, Appendix 2 |
| Information sources | 6 | Specify all databases, registers, websites, organisations, reference lists and other sources searched or consulted to identify studies. Specify the date when each source was last searched or consulted. | 4-5 |
| Search strategy | 7 | Present the full search strategies for all databases, registers and websites, including any filters and limits used. | Appendix 3 |
| Selection process | 8 | Specify the methods used to decide whether a study met the inclusion criteria of the review, including how many reviewers screened each record and each report retrieved, whether they worked independently, and if applicable, details of automation tools used in the process. | 5, Table 1 |
| Data collection process | 9 | Specify the methods used to collect data from reports, including how many reviewers collected data from each report, whether they worked independently, any processes for obtaining or confirming data from study investigators, and if applicable, details of automation tools used in the process. | 5 |
| Data items | 10a | List and define all outcomes for which data were sought. Specify whether all results that were compatible with each outcome domain in each study were sought (e.g. for all measures, time points, analyses), and if not, the methods used to decide which results to collect. | 4, Table 1 |
| | 10b | List and define all other variables for which data were sought (e.g. participant and intervention characteristics, funding sources). Describe any assumptions made about any missing or unclear information. | 4-5, Table 1 |
| Study risk of bias assessment | 11 | Specify the methods used to assess risk of bias in the included studies, including details of the tool(s) used, how many reviewers assessed each study and whether they worked independently, and if applicable, details of automation tools used in the process. | 5 |
| Effect measures | 12 | Specify for each outcome the effect measure(s) (e.g. risk ratio, mean difference) used in the synthesis or presentation of results. | 4-5, Table 1 |
| Synthesis methods | 13a | Describe the processes used to decide which studies were eligible for each synthesis (e.g. tabulating the study intervention characteristics and comparing against the planned groups for each synthesis (item #5)). | 5 |

| Section and Topic | Item # | Checklist item | Location where item is reported |
|---|---|---|---|
| | 13b | Describe any methods required to prepare the data for presentation or synthesis, such as handling of missing summary statistics, or data conversions. | N/A |
| | 13c | Describe any methods used to tabulate or visually display results of individual studies and syntheses. | 4-5 |
| | 13d | Describe any methods used to synthesize results and provide a rationale for the choice(s). If meta-analysis was performed, describe the model(s), method(s) to identify the presence and extent of statistical heterogeneity, and software package(s) used. | 4-5, 15-16 |
| | 13e | Describe any methods used to explore possible causes of heterogeneity among study results (e.g. subgroup analysis, meta-regression). | N/A |
| | 13f | Describe any sensitivity analyses conducted to assess robustness of the synthesized results. | N/A |
| Reporting bias assessment | 14 | Describe any methods used to assess risk of bias due to missing results in a synthesis (arising from reporting biases). | 4-5, 15-16 |
| Certainty assessment | 15 | Describe any methods used to assess certainty (or confidence) in the body of evidence for an outcome. | N/A |
| **RESULTS** | | | |
| Study selection | 16a | Describe the results of the search and selection process, from the number of records identified in the search to the number of studies included in the review, ideally using a flow diagram. | Figure 2 |
| | 16b | Cite studies that might appear to meet the inclusion criteria, but which were excluded, and explain why they were excluded. | N/A |
| Study characteristics | 17 | Cite each included study and present its characteristics. | Throughout the results section; Appendix 4 |
| Risk of bias in studies | 18 | Present assessments of risk of bias for each included study. | Appendix 5 |
| Results of individual studies | 19 | For all outcomes, present, for each study: (a) summary statistics for each group (where appropriate) and (b) an effect estimate and its precision (e.g. confidence/credible interval), ideally using structured tables or plots. | N/A – narrative synthesis. Individual study outcomes presented in Appendix 4. |
| Results of syntheses | 20a | For each synthesis, briefly summarise the characteristics and risk of bias among contributing studies. | Throughout the results section, Appendix 4, Appendix 5 |
| | 20b | Present results of all statistical syntheses conducted. If meta-analysis was done, present for each the summary estimate and its precision (e.g. confidence/credible interval) and measures of statistical heterogeneity. If comparing groups, describe the direction of the effect. | N/A |
| | 20c | Present results of all investigations of possible causes of heterogeneity among study results. | N/A |
| | 20d | Present results of all sensitivity analyses conducted to assess the robustness of the synthesized results. | N/A |
| Reporting | 21 | Present assessments of risk of bias due to missing results (arising from reporting biases) for each synthesis | N/A |

| Section and Topic | Item # | Checklist item | Location where item is reported |
|---|---|---|---|
| biases | | assessed. | |
| Certainty of evidence | 22 | Present assessments of certainty (or confidence) in the body of evidence for each outcome assessed. | N/A |
| **DISCUSSION** | | | |
| Discussion | 23a | Provide a general interpretation of the results in the context of other evidence. | 13-15 |
| | 23b | Discuss any limitations of the evidence included in the review. | 15-16 |
| | 23c | Discuss any limitations of the review processes used. | 15-16 |
| | 23d | Discuss implications of the results for practice, policy, and future research. | 16 |
| **OTHER INFORMATION** | | | |
| Registration and protocol | 24a | Provide registration information for the review, including register name and registration number, or state that the review was not registered. | 4 |
| | 24b | Indicate where the review protocol can be accessed, or state that a protocol was not prepared. | 4 |
| | 24c | Describe and explain any amendments to information provided at registration or in the protocol. | N/A |
| Support | 25 | Describe sources of financial or non-financial support for the review, and the role of the funders or sponsors in the review. | 17 |
| Competing interests | 26 | Declare any competing interests of review authors. | 17 |
| Availability of data, code and other materials | 27 | Report which of the following are publicly available and where they can be found: template data collection forms; data extracted from included studies; data used for all analyses; analytic code; any other materials used in the review. | Appendix 4, Appendix 5 |

*From:* Page MJ, McKenzie JE, Bossuyt PM, Boutron I, Hoffmann TC, Mulrow CD, et al. The PRISMA 2020 statement: an updated guideline for reporting systematic reviews. BMJ 2021;372:n71. doi: 10.1136/bmj.n71

For more information, visit: http://www.prisma-statement.org/
