## Supplementary materials 2 - appendix 4 for "Strengthening vaccination delivery system resilience in the context of protracted humanitarian crisis: a realist-informed systematic review"

| | | | | | | Mechanism | | | | | | | | | | | | |
|---|---|---|---|---|---|---|---|---|---|---|---|---|---|---|---|---|---|---|
| | | | | | | | Activities: System "hardware" ↔ System "software" | | | | | | | | | | | |
| **First author** | **Intervention class** | **Country** | **Antigen** | **Study design** | **Context** | *Type* | *Increased material resource availability* | *Increased human resource availability and capability* | *Changes to collaterals and redundancy* | *Changes to networks and collaboration* | *Increased flexibility (including decentralisation)* | *Improved situational awareness and information management* | *Improved preparedness and planning* | *Strengthened leadership* | *Changes to organisational and wider system culture* | *Improvement population trust* | *Description* | **Outcome(s)** |
| Amani (34) | Campaign | Cameroon | Cholera | Program evaluation | Epidemic response focused on South/South-West of the country in context of insecurity, political crisis, and significant constraints to health service availability, access to WASH and potable drinking water, and livelihood-linked population movements. | Adaptive | | | | | | | | | | | Reactive intervention following mobilisation of vaccine doses from the ICG stockpile. Multi-pronged campaign under oversight of Cameroonian Ministry of Health, involving recruitment and training of vaccinators and social mobilisers. Heavy emphasis on community mobilisation through public messaging, advocacy with religious and community leaders to improve both awareness and trust among affected populations. | Overall post-campaign vaccination coverage of around 82% but with large geographical and age-based variations (>95% among adolescents in the intervention area, <80% in adults aged >20yrs. |
| Amani (35) | Campaign | Cameroon | Meningitis | Program evaluation | Conflict-affected setting. Focus on refugee camps in the far North and East of the country, in the context of population movement from CAR, Chad and Nigeria. Existing health service access in affected areas predominantly supported by international relief agencies. | Adaptive | | | | | | | | | | | Preventive mass campaign against Men ACWY targeting children aged 2+, under coordination of national Ministry of Health with international agency and donor financial support. Recruitment and training of vaccinators and community mobilisers. Interactions with religious, community and other camp leaders to increase uptake through improved awareness and trust in service providers. | Overall coverage post-campaign was 81.97% but with large variations by geography and by age, and only two areas achieved the target coverage of >95%. Impact of campaign affected by (i) ongoing population movement into affected areas, and difficulties in tracking new arrivals to ensure vaccination, and (ii) limitations to cold chain capacity given unreliable power supplies. |
| Aradhi (67) | Health information and surveillance | Iraq | Polio | Program evaluation | Conflict-affected setting. Reports surveillance and immunisation response in Karbala province to confirmed polio cases identified in Baghdad in 2014. Wider health system context in Iraq at this time was influenced by ongoing conflict and included destruction of health facilities, health worker attrition and resulting access constraints, as well financial resourcing shortfalls. | Adaptive | | | | | | | | | | | Multi-pronged response including SIAs, household outreach, in-person surveillance work to follow-up on reported AFP cases at facility level, among other activities, to help target vaccination and improve awareness of the importance of immunisation among affected populations. | 11 AFP cases were reported from Karbala during the course of the study. Increases in vaccination coverage for OPV3, BCG and measles vaccination were recorded in Karbala following the intervention. |

| | | | | | | Mechanism | | | | | | | | | | | | |
|---|---|---|---|---|---|---|---|---|---|---|---|---|---|---|---|---|---|---|
| | | | | | | | Activities: System "hardware" ↔ System "software" | | | | | | | | | | | |
| **First author** | **Intervention class** | **Country** | **Antigen** | **Study design** | **Context** | *Type* | *Increased material resource availability* | *Increased human resource availability and capability* | *Changes to collaterals and redundancy* | *Changes to networks and collaboration* | *Increased flexibility (including decentralisation)* | *Improved situational awareness and information management* | *Improved preparedness and planning* | *Strengthened leadership* | *Changes to organisational and wider system culture* | *Improvement population trust* | *Description* | **Outcome(s)** |
| Arale (62) | Governance and coordination | Multiple | Polio | Program evaluation | Complex humanitarian crisis. Intervention addressing polio prevention and control in Kenya and Somalia in context of porous borders, movement of displaced and nomadic populations, insecurity, and limited health service access due to geographical remoteness of locations and limited resourcing from the centre (low funding, workforce recruitment and retention difficulties, low skills base of health workers). | Transfor-mative | | | | | | | | | | | Multi-dimensional governance and coordination intervention involving establishment of new cross-border coordination mechanisms, and governance bodies in Kenya and Somalia bringing together national and local partners; harmonisation of recruitment approaches for community health volunteers, and information exchange (on informal border crossing points, migratory routes etc); capacity building for community-based surveillance activities, and other approaches. These activities contributed to improved situational awareness (including hotspot identification) to support vaccination targeting, and improved service delivery through training and standardisation. | Large year-on-year increases in the number of children <5 vaccinated through SIAs linked to the programme, in the number of children aged 12-59 months in receipt of at least 1 dose of polio vaccine, and in the number of AFP cases reported at district level (due to improved case reporting). Progressive increases over the lifecycle of the intervention in the number of community health volunteers recruited, cross-border initiative coordination meetings held, and other intermediate measures. |
| Baptiste (33) | Campaign | Nigeria | Measles | Other | Complex humanitarian crisis. Campaign carried out in northern Nigerian Borno State in context of large, national measles outbreak, and targeting mixed population of IDPs, refugees and nomads living in and outside camp settings, but all with limited access to health services through fixed sites. Wider context of ongoing insecurity in Borno linked to the Boko Haram insurgency. | Adaptive | | | | | | | | | | | Reactive mass vaccination campaign carried out in two phases under coordination of Nigerian government. Describes use of polio eradication architecture including case-based measles surveillance that is integrated with AFP surveillance for identification of hotspots, supported by newly trained community health workers. Hard-to-reach settlements were accessed with support of military escorts (in event of insecurity) and use of local infrastructure through the GPEI, contributing to improved service reach. | Reported weighted coverage of 85.7% post-campaign (95%CI: 79.6–90.1). Reported coverage was highest in children under 9 months, and 24-35 months at 100% and 92.8% respectively. Study authors note difficulties in coverage estimation in context of ongoing population movement in affected areas. |
| Bawa (58) | Service integration | Nigeria | Multiple | Program evaluation | Conflict-affected setting. Service changes in response to polio cases reported in Northern States (Bauchi, Borno, Kano and Yobe) in context of ongoing insecurity linked to insurgency, and long-term service access challenges resulting from the challenging terrain, and ongoing population movement (nomadic and forced). | Adaptive | | | | | | | | | | | Expanded implementation of existing intervention offering routine immunization together with a basic integrated package of primary healthcare interventions focused on maternal, newborn and child health. This was coordinated through the national polio emergency operations centre, with financial and logistical support from international actors (principally WHO, UNICEF and the BMGF). Use of mobile health teams to reach outlying areas, and monitoring of reach using geographical information tracking. Local engagement through community mobilisers, recruited from affected communities. | At baseline, 19.6% of children in the surveyed intervention area had routine immunisation cards 17.8% were fully immunized. By the end of the survey period, 49.1% had routine immunisation cards and 49.0% were fully immunized. The proportion of zero dose children declined from 11.5% to 4.7%. |

| First author | Intervention class | Country | Antigen | Study design | Context | Mechanism | | | | | | | | | | | | Outcome(s) |
|---|---|---|---|---|---|---|---|---|---|---|---|---|---|---|---|---|---|---|
| | | | | | | Type | Activities: System "hardware" ↔ System "software" | | | | | | | | | | Description | |
| | | | | | | | Increased material resource availability | Increased human resource availability and capability | Changes to collaterals and redundancy | Changes to networks and collaboration | Increased flexibility (including decentralisation) | Improved situational awareness and information management | Improved preparedness and planning | Strengthened leadership | Changes to organisational and wider system culture | Improvement population trust | | |
| Bonfrer (50) | Health financing | Burundi | Multiple | Time series analysis | Conflict-affected setting. At the time of the study Burundi was among the lowest income countries in the world with a GDP of 251 current US$ per capita, and a health system recovering from civil war. Study focused on implementation of an adaptive intervention at national level. | Adaptive | | | | | | | | | | | Intervention acting through changes to facility-level incentive structures to enhance supply of services for various interventions including childhood vaccination. Implementation of performance-based financing, with around 700 health care facilities nationally paid retrospectively based on the quantity and quality of services provided. Bundled services included antenatal care, HIV preventive interventions and delivery of routine immunisations to children under 1 year of age. | Statistically significant increase in uptake of a full vaccination package, including all of its components (BCG, polio, DTP and measles) following PBF introduction, especially among the poorest households. |
| Bwire (46) | Campaign | Uganda | Cholera | Cross-sectional survey | Complex humanitarian crisis linked primarily to population displacement from neighbouring DRC. Response concerned with refugees living inside and outside refugee camps, but also to address risk of infection spread to host communities in affected areas of northern Uganda. | Adaptive | | | | | | | | | | | Reactive, multi-pronged intervention led by Ugandan Ministry of Health, initially incorporating WASH interventions and enhanced surveillance following refugee movement, but then including a vaccination campaign delivered in partnership with WHO, UNICEF and MSF among other partners once the first cholera cases had been identified. Campaign delivered as intensive intervention to 390,000 households in four targeted areas, using OCV, using a door-to-door approach to improve service access. | Estimated coverage of 94% (95% CI 92-95%) with at least one dose, and 78% (76-81%) with two OCV doses following the campaign. |
| Dadari (47) | Health financing | Multiple | Multiple | Program evaluation | Multiple settings – study evaluates Gavi-supported, health system strengthening programming in 13 countries, including a number meeting the inclusion criteria for the study (Afghanistan, CAR, DRC for example). Large variations in context according to the country concerned. | Adaptive | | | | | | | | | | | Health system strengthening activities to promote equity in vaccination delivery across populations – focusing specifically on the use of pro-equity strategies to improve governance. Multi-pronged approaches incorporating strengthened situational awareness systems, human resource capacity building (e.g. peer-support groups for health workers in remote areas), mobilisation efforts using community outreach volunteers and local community champions to improve uptake. | Marginal improvements in national vaccination coverage trends for DTP3 in countries eligible for inclusion in this review over the period 2016-19 (patterns variable by country). Primary outcome focus of the study was on implementation of pro-equity strategies in Gavi partner countries. |

| | | | | | | Mechanism | | | | | | | | | | | | |
|---|---|---|---|---|---|---|---|---|---|---|---|---|---|---|---|---|---|---|
| | | | | | | | Activities: System "hardware" ↔ System "software" | | | | | | | | | | | |
| ***First author*** | ***Intervention class*** | ***Country*** | ***Antigen*** | ***Study design*** | ***Context*** | *Type* | *Increased material resource availability* | *Increased human resource availability and capability* | *Changes to collaterals and redundancy* | *Changes to networks and collaboration* | *Increased flexibility (including decentralisation)* | *Improved situational awareness and information management* | *Improved preparedness and planning* | *Strengthened leadership* | *Changes to organisational and wider system culture* | *Improvement population trust* | *Description* | ***Outcome(s)*** |
| Dadgar (40) | Campaign | Afghanistan | Measles | Cross-sectional survey | Conflict-affected setting. Study focused on the period 2001-2, shortly after Coalition operations began. The campaign described continued one that had begun under the Taliban administration but was disrupted by the invasion, on a background of low overall life expectancy, 25% mortality in children under 5, and poor access to primary healthcare services including in the areas of central Afghanistan addressed in this study. | Adaptive | | | | | | | | | | | Preventive and reactive vaccination campaign targeting children aged 6mo-12years in central Afghanistan in light of high background measles incidence rates, known low coverage and an ongoing outbreak over the preceding 2 years. Intervention built on architecture previously developed for National Immunisation Days. Children were targeted for vaccination irrespective of prior vaccination status. Campaign coordinated by Ministry of Health with international support from WHO, UNICEF and national and international NGOs. Delivery supported by health worker training and task shifting, use of mobile teams, and administration in community settings including mosques. | 77% of children in targeted areas identified as eligible were vaccinated, with reported coverage being lowest in the capital, Kabul (62%). Estimates influenced by significant difficulties in denominator estimation especially in Kabul, where population movement (in and out) had been high in the months leading up to, and following, the invasion. |
| Duru (63) | Health workforce | Nigeria | Polio | Program evaluation | Conflict-affected setting. Intervention focused on northern Nigerian states affected by chronic insecurity, but also low health service access, health worker shortages, persistently low vaccination coverage and intermittently reported polio cases. Broader social context affecting vaccination uptake including distrust of public health providers, and religious factors. | Adaptive | | | | | | | | | | | Introduction and use of Volunteer Community Mobilisers (VCMs) to support polio vaccination delivery. Supported by multi-level supervision arrangements, VCMs were used to support surveillance through soft intelligence gathering, training to support vaccine logistics and monitoring, and worked with communities (from which they were recruited in the first place) to promote uptake, in partnership with local religious and community leaders. | Principal focus of the intervention was on reducing the number of zero dose children through reductions in missed opportunities for vaccination. Study reports decline in the percentage of missed children, from 4.5% in 2014 to 0.8% in 2018. |
| Edmond (54) | Service integration | Afghanistan | Multiple | Case-control study | Conflict-affected setting. Describes integrated service delivery in 54 intervention and control districts in central, southern and northern Afghanistan in context of ongoing conflict, poor access to health services, and continuing access challenges, with less than 50% of infants in 2015 in receipt of all vaccinations per the EPI schedule. | Adaptive | | | | | | | | | | | Expanded use of Mobile Health Teams (MHTs), coordinated through Ministry of Health. MHTs offered services in accordance with the basic package of care agreed nationally, visiting each village within their allocated geographical area intermittently for 1-2 days at a time to deliver primary care services. Services delivered were recorded in electronic health information systems. MHTs had fixed structures, and health workers received induction and annual refresher training. | Outcome measurement focused on vaccine visits as proxies for uptake. Observed effects differed by antigen. Pentavalent and measles vaccine visits were similar in clinics in the intervention and control districts overall, but there was a statistically significant improvement in receipt of first dose measles vaccine in the intervention sites compared to controls (p=0.02). |

| *First author* | *Intervention class* | *Country* | *Antigen* | *Study design* | *Context* | *Mechanism* | | | | | | | | | | | | *Outcome(s)* |
|---|---|---|---|---|---|---|---|---|---|---|---|---|---|---|---|---|---|---|
| | | | | | | *Type* | *Activities: System "hardware" ↔ System "software"* | | | | | | | | | | *Description* | |
| | | | | | | | *Increased material resource availability* | *Increased human resource availability and capability* | *Changes to collaterals and redundancy* | *Changes to networks and collaboration* | *Increased flexibility (including decentralisation)* | *Improved situational awareness and information management* | *Improved preparedness and planning* | *Strengthened leadership* | *Changes to organisational and wider system culture* | *Improvement population trust* | | |
| Elsayed (36) | Campaign | Sudan | Measles | Other | Complex humanitarian crisis. Focus on Darfur in Sudan in context of armed conflict over the preceding year, internal displacement and refugee movement affecting >1m people in Sudan and into neighbouring Chad; limited access to WASH, nutrition and basic health services among displaced populations, and measles transmission noted among IDPs in camps in the area. | Adaptive | ■ | ■ | ■ | ■ | | | | | | ■ | Reactive vaccination campaign under coordination of Ministry of Health, working with WHO, UNICEF and other partners, but with support from a technical working group including NGOs. The campaign was donor funded, and depended for success on prior negotiation of physical access with parties to the ongoing conflict. Mixed mode of delivery combining fixed sites, temporary posts and mobile outreach. Service delivery integrated with polio vaccination and nutritional supplementation given needs in the target populations. Vaccination activities supported by community mobilisation via mass media and community level activities to bolster demand. | Around 93% of the accessible population (defined on the basis of residence in areas where security allowed a higher degree of access) and 77% of the total target population were vaccinated during the campaign. Coverage was highest in most secure areas (South Darfur), but lowest in parts of the country (West Darfur) where accessibility was most challenging because of security, terrain and other factors. |
| Engineer (52) | Health financing | Afghanistan | Multiple | RCT | Conflict-affected setting. Introduction of payment for performance (P4P) across 11 provinces in 442 primary care facility level in Afghanistan, in the context of ongoing conflict and instability. Authors note prior experience in Afghanistan in use of P4P to support aspects of contracting for the basic health service package. | Adaptive | ■ | ■ | | | | | | | | | Introduction of P4P, with bonuses paid to healthcare workers, based on quality scoring for services delivered linked to information gathered via the HMIS. Health facilities also submitted monthly activity reports that were used to inform the size of incentive payments that were made. Managing NGOs retained 10% of the payments centrally. Mode of payment to health workers themselves was determined by facility managers (e.g. blanket or weighted payments) | Similar effects were observed at end-point by comparison with baseline, although for one antigen (pentavalent vaccination) coverage in the pre-post period compared with the comparison areas actually declined (−5.7%, P 0.01). Speculated reasons for limited effects of this intervention include the timeliness of payment to health workers (which was not optimal) and incentivisation that was insufficient to promote the desired performance improvements. |
| Falisse (51) | Health financing | Burundi | Multiple | Time series analysis | Conflict-affected setting. Describes wider roll out of Burundi in post-conflict period, following an initial period of piloting in 2006-10. Wider health system context was one of chronic under-funded and human resource attrition partly linked to insecurity but also low pay and poor working conditions. | Adaptive | ■ | | | ■ | | | | | | | Wider roll out of P4P through facility-level contracting. Intervention involves subsidy payments to facilities based on the volume of services given (but without an explicit service quality premium attached to these payments). The intervention was delivered with funding support from the EU and external technical support, under leadership of national Ministry of Health. | No statistically significant effect in vaccination uptake by antigen (BCG, polio, MMR, DTP) or overall in children in the intervention areas following introduction of P4P. |

| | | | | | | ***Mechanism*** | | | | | | | | | | | | |
|---|---|---|---|---|---|---|---|---|---|---|---|---|---|---|---|---|---|---|
| | | | | | | | *Activities* | | | | | | | | | | | |
| | | | | | | | *System "hardware"* ↔ *System "software"* | | | | | | | | | | | |
| ***First author*** | ***Intervention class*** | ***Country*** | ***Antigen*** | ***Study design*** | ***Context*** | *Type* | *Increased material resource availability* | *Increased human resource availability and capability* | *Changes to collaterals and redundancy* | *Changes to networks and collaboration* | *Increased flexibility (including decentralisation)* | *Improved situational awareness and information management* | *Improved preparedness and planning* | *Strengthened leadership* | *Changes to organisational and wider system culture* | *Improvement population trust* | *Description* | ***Outcome(s)*** |
| Fekadu (60) | Governance and coordination | Angola | Polio | Program evaluation | Conflict-affected setting. Intervention focuses on efforts to reduce risk of polio transmission in the early post-conflict phase, against background of poor accessibility in affected districts due to insecurity and poor infrastructure. In 1999, Angola had experienced the biggest poliomyelitis epidemic ever recorded in the African Region up to that point, with 1117 cases and 113 deaths caused by wild poliovirus type 1 and 3. | Transfor-mative | | | | | | | | | | | Partnership intervention with the Angolan military to support polio vaccination delivery and integrated services in security compromised areas, creating wholly new service delivery architecture. Activities were coordinated by an inter-agency coordination committee under the Ministry of Health, but on which military representatives also sat. Included support to administration of oral polio vaccine, vitamin A, deworming agents, social mobilization, monitoring campaign quality, surveillance and logistics. Military vaccinators were also engaged in community mobilisation activities, mostly through information provision (leafleting etc). | Presents soft data on outcomes only. Reports that "the average number of children vaccinated per day by a military vaccination team was found to be double than the other volunteer vaccinators. It was also witnessed that there were less number of missed children in military covered area than the areas covered by vaccination volunteers". |
| Habib (59) | Service integration | Pakistan | Polio | RCT | Conflict-affected setting. Integrated polio vaccination and prevention activities delivered in geographical areas specifically chosen for service delivery challenges linked to ongoing insurgency (Bajaur and Karachi) and general insecurity (Kashmore). Wider context of ongoing polio endemicity in Pakistan at the time of the trial, in common with only a handful of other countries globally at this time. | Transfor-mative | | | | | | | | | | | Two intervention arms offered vaccine (OPV or IPV depending on the arm) in addition to comprehensive community mobilisation activities, enhanced communication work, and multiple service delivery modes combining fixed vaccination points as well as mobile "camps". Delivery was via teams comprising both vaccinators and community outreach workers. Prior negotiation with community leaders also occurred to ensure access. | The proportion of fully vaccinated children increased in the two intervention arms compared with the control arm (7.3% [95% CI 4.5–10.0] increase in the first intervention arm vs control; 9.5% [6.9–12.0] increase in second intervention arm vs control; overall p<0·0001). |
| Haddison (78) | Multi-dimensional | Cameroon | Polio | Program evaluation | Conflict-affected setting. Focus of the intervention in far north of the country, against background of insecurity linked to Boko Haram insurgency, and then new political unrest in the northwest and southwest. Health facilities in the affected areas closed for periods owing to insecurity, leading to disruptions to routine immunisation delivery. | Adaptive | | | | | | | | | | | Combined "Mother and Child Week" and SIA with overall objective of improving polio vaccination coverage in affected areas, coordinated through an existing central technical group for the EPI programme. Funds were drawn from the Ministry of Health, WHO and UNICEF. Focus on recruitment and training of vaccination workers, and delivery through combination of fixed sites and mobile outreach. Supported by community mobilisation activities including engagement with local religious and community leaders. | Reported administration coverage following the SIA of 89.9% compared to 91.2% following a preceding campaign in 2017 (although data for the earlier campaign are disputed owing to problems with denominator estimation). Notes variations in reported coverage by geographical area and a number of administrative estimates in excess of 100% suggesting inaccurate denominator estimation. |

| | | | | | | Mechanism | | | | | | | | | | | | |
|---|---|---|---|---|---|---|---|---|---|---|---|---|---|---|---|---|---|---|
| | | | | | | | Activities: System "hardware" ↔ System "software" | | | | | | | | | | | |
| **First author** | **Intervention class** | **Country** | **Antigen** | **Study design** | **Context** | *Type* | *Increased material resource availability* | *Increased human resource availability and capability* | *Changes to collaterals and redundancy* | *Changes to networks and collaboration* | *Increased flexibility (including decentralisation)* | *Improved situational awareness and information management* | *Improved preparedness and planning* | *Strengthened leadership* | *Changes to organisational and wider system culture* | *Improvement population trust* | *Description* | **Outcome(s)** |
| Hamman-yero (70) | Community engagement and mobilisation | Nigeria | Multiple | Program evaluation | Conflict-affected setting. Describes community engagement activities in four northern Nigerian states (Kano, Bauchi, Borno, and Yobe), some affected by chronic insecurity. | Adaptive | | | | | | | | | | | Community engagement and mobilisation intervention bolstering training for existing health service personnel, introducing supervisory checklists for community outreach activities, and working with community and religious leaders to sensitive communities prior to mobile health team visits. Volunteer mobilisers were used to help refer children and pregnant women into services to improve uptake. | OPV coverage for children below 1 year of age was 44% at baseline; this rose to 76% overall by the fourth quarter of the intervention. Large variations in coverage across the geographical areas for the intervention were noted however. Some variations were noted also according to the antigen – third dose pentavalent vaccine coverage rose from 22% to 62% in eligible children in the intervention period over the four quarters. |
| Idris (57) | Service integration | South Sudan | Multiple | Program evaluation | Complex humanitarian crisis. Intervention implementation in South Sudan in context of ongoing conflict, population movement and high levels of underlying population vulnerability including malnutrition, limited health service access. Prior to the intervention, immunisation services had predominantly been delivered in a vertical way linked to specific programme funding. | Adaptive | | | | | | | | | | | Integration of immunisation and nutrition services in one arm of the intervention, and immunisation with regular paediatric outpatient services in the other arm (these arms were geographically separated by county boundaries). Intervention implementation was supported by a decentralised governance model allowing a degree of discretion for local service managers in how to integrate. The paediatric outpatient department intervention also included use of uptake-promotion measures such as service audits and calendar reminders to patients. | Increases in uptake by antigen were seen for all vaccines delivered across both the nutrition-immunisation service integration sites, and the outpatient-immunisation service integration sites. This effect was more pronounced for first dose pentavalent vaccine in nutrition-immunisation sites that in outpatient-immunisation sites (rate ratio of 1.23 (95% CI 1.12-1.36) but no significant difference was seen for other doses or antigens. Statistically significant improvements in uptake across a range of antigens and doses were noted in nutrition-immunisation integration sites by comparison with baseline. |
| Kamadjeu (72) | Multi-dimensional | Somalia | Polio | Program evaluation | Complex humanitarian crisis featuring ongoing conflict, population displacement and significant constraints on health service access. Background to the intervention was a large outbreak (189 confirmed WPV1 cases) of polio in Mogadishu and surrounding districts, and active attempts by actors opposed to the government to ban immunisation among populations living under their jurisdiction. | Adaptive | | | | | | | | | | | SIAs launched to improve OPV uptake among children <5, children <10 and then people of any age successively. Cross-border coordination was supported by coordination meetings in Somalia and in Kenya. Service delivery was via a combination of fixed posts and mobile outreach. This was supported by intensive community engagement to bolster uptake. | Overall coverage of OPV across assessed districts following the campaign varied from 72% to 82%, from a national baseline of <50% in 2012 (prior to the outbreak). By the end of 2013 there had been an unbroken period of 25 weeks with no new WPV cases reported – which the study authors took as indicative of the success of the SIAs. |
| Khan (38) | Campaign | Bangladesh | Multiple | Cross-sectional survey | Complex humanitarian crisis. Focused on displaced Rohingya refugees from neighbouring Myanmar, on background of ongoing conflict, and temporary residence for up to a million refugees in 32 camps scattered across Cox's Bazaar. Most refugees at the time of the study were living in makeshift shelters with limited access to WASH and health interventions. | Adaptive | | | | | | | | | | | Vaccination campaign spanning measles, rubella, poliomyelitis and oral cholera vaccine for all individuals living in Cox's Bazaar save for children under the age of 1. The target population group for poliomyelitis vaccination was children aged 0-5. | Reported coverage post-campaign varied from as low as 38% (measles-rubella) to 94% for first dose oral cholera vaccine. Possible reasons for this variation identified by the study team included (i) the short time span for the campaign; (ii) continuing population displacement into Cox's Bazaar during the campaign; and (iii) low awareness amongst the refugee population. |

| | | | | | | ***Mechanism*** | | | | | | | | | | | | |
|---|---|---|---|---|---|---|---|---|---|---|---|---|---|---|---|---|---|---|
| | | | | | | | *Activities* — *System "hardware"* ↔ *System "software"* | | | | | | | | | | | |
| ***First author*** | ***Intervention class*** | ***Country*** | ***Antigen*** | ***Study design*** | ***Context*** | *Type* | *Increased material resource availability* | *Increased human resource availability and capability* | *Changes to collaterals and redundancy* | *Changes to networks and collaboration* | *Increased flexibility (including decentralisation)* | *Improved situational awareness and information management* | *Improved preparedness and planning* | *Strengthened leadership* | *Changes to organisational and wider system culture* | *Improvement population trust* | *Description* | ***Outcome(s)*** |
| Khetsuriani (73) | Multi-dimensional | Ukraine | Polio | Program evaluation | Conflict-affected setting. Broader context of conflict with Russia (2014) and ongoing political and economic crisis, as well as large declines in coverage across a range of antigens observed since 2009 owing to (i) rising vaccine hesitancy, (ii) declining resource allocations to health in face of ongoing crisis, and (iii) low acceptance among health care workers among other factors. | Adaptive | | | | | | | | | | | Multi-dimensional intervention focused on SIAs but supported by a new national governance mechanism bringing together the Ukrainian government with international partners including WHO and UNICEF to support polio vaccination delivery. International partners provided vaccines, technical, financial and logistical support and helped develop advocacy materials to support the SIAs. Wider capacity building included training activities for HCWs but also for journalists to try to improve public messaging, and was supported by wider advocacy and community engagement activities. | Reported OPV coverage increased from 64.4% following the first SIA round to 80.7% following round 3. Effects on public perceptions of the importance of polio vaccination were also documented: awareness of polio among caregivers increased from 68% at the beginning of the outbreak to 89% following SIA round 1, 91% following round 2, and 96% following round 3, mirrored by rising support for the need for supplemental polio vaccination among these populations. |
| Korir (71) | Community engagement and mobilisation | Nigeria | Polio | Time series analysis | Conflict-affected setting. Intervention implementation in a series of northern Nigerian states including Borno, which at the time of the study was subject to ongoing, insurgency-related insecurity. The broader operating context in Borno at this time included poor access to health services and health worker attrition owing to a range of factors including low pay. | Adaptive | | | | | | | | | | | Micro-level intervention to promote directly observed polio vaccination, accompanied by incentives to draw parents into services to have their children vaccinated. In Borno specifically, members of a military liaison force helped provide security to support vaccination delivery in outlying areas. Incentives to parents were in kind (soap, milk sachets, noodles, sugar etc) and were supported by community engagement activities with local leaders. | Overall decline in the number of zero-dose children across all targeted geographical areas from 2.4% in August 2014 (baseline) to 1.1% in May 2016 (endpoint). Steady appreciation in the proportion of children in receipt of four doses or more of OPV in Borno from 75% in 2013 to 86% in 2016. After a cluster of vaccine-derived polio cases in Borno in 2014 prior to the intervention, 1 further case was reported over the following 24 months. |
| Lam (41) | Campaign | Iraq | Cholera | Cross-sectional survey | Complex humanitarian crisis. Cholera outbreak in the context of ongoing conflict and high levels of internal displacement in Iraq, and refugee arrivals from neighbouring Syria – the intervention described focused on IDP camps. Displacement settings were frequently overcrowded and with poor access to shelter, WASH and basic health services. The wider health system context in Iraq during this period included financial austerity measures linked to an ongoing economic crisis, which reduced resources for front-line services and contributed to ongoing problems with cholera transmission. | Adaptive | | | | | | | | | | | Reactive campaign using OCV. A 2-dose campaign was designed targeting >255,000 aged 1 year and over living in IDP camps, refugee camps and other collective settings. Campaign coordination was provided by the Iraqi Ministry of Health working in partnership with international and local actors. Delivery was via a combination of fixed sites and door-to-door outreach and used vaccination teams of fixed composition (1 vaccinator, 1 recorder and 1 "crowd controller"), and included the use of community mobilisers and public information campaigns to draw service users in, although only 10% of those surveyed reported having received any information about the campaign via these routes. | 2-dose OCV coverage in the targeted camps was reported as 87% (95% CI 85%–89%). Two-dose OCV coverage in 3 northern governorates targeted (91%; 87%–94%) was higher than that in the 7 southern and central governorates (80%; 77%–82%). |

| First author | Intervention class | Country | Antigen | Study design | Context | Mechanism: Type | Activities (System "hardware" ⟷ System "software"): Increased material resource availability | Increased human resource availability and capability | Changes to collaterals and redundancy | Changes to networks and collaboration | Increased flexibility (including decentralisation) | Improved situational awareness and information management | Improved preparedness and planning | Strengthened leadership | Changes to organisational and wider system culture | Improvement population trust | Description | Outcome(s) |
|---|---|---|---|---|---|---|---|---|---|---|---|---|---|---|---|---|---|---|
| Lubogo (39) | Campaign | Somalia | Cholera | Cross-sectional survey | Complex humanitarian crisis. Reactive campaign implemented in context of ongoing conflict, high levels of internal displacement, and in the specific time period of this outbreak, a drought contributing to depleted water sources and further concentration of IDPs in settlements with limited access to WASH and basic health services. | Adaptive | ■ | ■ | | | | ■ | | | | ■ | Reactive, two-round vaccination campaign implemented in 11 targeted districts in 2017. The mode of service delivery was via a combination of fixed sites (including schools, health services and public spaces) and door-to-door outreach, to maximise reach, and using a phased approach to focus efforts on a specific district before moving on to the next one. The campaign was preceded by a microplanning exercise led by the Ministry of Health, with international partner support, and included training for vaccinators, as well as the use of community mobilisers (selected for prior experience through polio eradication work) to promote uptake. | Reported coverage for two doses of OCV following the campaign was 92.5%, with 7.0% receiving just a single dose. There were large variations in coverage by district, ranging from a maximum of 100%, to as low as 85.9% in one area. |
| Mbaeyi (74) | Multi-dimensional | Multiple | Polio | Program evaluation | Complex humanitarian crisis. Describes regional response to a polio outbreak focused on Syria and Iraq, in context of ongoing conflicts in both countries, large cross-border movements and high levels of internal displacement. This occurred against a wider health system context of financial resource constraints, health worker flight and outright destruction of health facilities (in Syria in particular), and broader governance fragmentation. | Transfor-mative | ■ | ■ | | ■ | | ■ | ■ | ■ | | ■ | Governments of eight MENA region countries designated the outbreak a public health emergency and worked with Global Polio Eradication Initiative (GPEI) to develop a regionally-coordinated response plan focused on improved surveillance for AFP cases, and polio vaccine administration to over 27m children regionally. The plan was multi-phase, focusing initially on halting transmission, then identifying importation and transmission hotspots, and finally immunisation system strengthening. Surveillance strengthening work included introduction of EWARN systems in some countries, and improvement to case detection. Access in areas of insecurity was secured through negotiation with community leaders. | Reported effects varied by country, with generally better evidence of impact in Syria than Iraq (of the two countries most directly affected by conflict). The proportion of non-polio AFP cases among children aged 6–59 months who were reported to have received at least 3 doses of OPV in Syria rose from 82% in 2013 to 94% in 2015, but was unchanged at 93% among Iraqi children of the same age during this period. Of those children aged 6–59 months with non-polio AFP, the proportion who had never received polio vaccination of any form decreased from 9% in 2013 to 2% in 2015 in Syria, but rose from 1% to 3% in Iraq over the same period. |

| | | | | | | *Mechanism* | | | | | | | | | | | | |
|---|---|---|---|---|---|---|---|---|---|---|---|---|---|---|---|---|---|---|
| | | | | | | | *Activities* — *System "hardware"* ↔ *System "software"* | | | | | | | | | | | |
| ***First author*** | ***Intervention class*** | ***Country*** | ***Antigen*** | ***Study design*** | ***Context*** | *Type* | *Increased material resource availability* | *Increased human resource availability and capability* | *Changes to collaterals and redundancy* | *Changes to networks and collaboration* | *Increased flexibility (including decentralisation)* | *Improved situational awareness and information management* | *Improved preparedness and planning* | *Strengthened leadership* | *Changes to organisational and wider system culture* | *Improvement population trust* | *Description* | ***Outcome(s)*** |
| Mbaeyi (75) | Multi-dimensional | Syria | Polio | Program evaluation | Conflict-affected setting. Describes response to a new polio outbreak in the context of ongoing Civil War in Syria, widespread destruction of health facilities, health worker attrition and steep declines in population coverage of key antigens since the war began. In addition, bans on vaccination campaigns had intermittently been imposed by governing authorities in the targeted governorates. In 2010, the national 3-dose OPV coverage estimate by the age of 1 83%, but by 2016 this had fallen to 48%. In addition, the success of previous OPV SIAs following a polio outbreak in 2013-14 appeared to have been limited: reported administrative coverage following these two campaigns was 7% and 23% respectively. | Adaptive | | | | | | | | | | | Reactive SIAs following identification of polio cases, with activities focused on two governorates (one of which was nominally under the control of Islamic State at the time of the first round). The campaign used OPV for all children <5, but IPV was added for the second round for children aged 2-23 months. Social mobilisation and community engagement activities to improve public perceptions of vaccination were a key mechanism for improving uptake among targeted populations. | Reported coverage improved following SIA rounds. Following round 1, coverage in one rose from 79% to 88%, but the change in the second was uncertain owing to large differences between administrative coverage and post-campaign monitoring estimates. Post-campaign monitoring results for the governorate most affected by conflict at the time (Raqqa) indicated a large increase in second dose OPV coverage from 57% in round 1, to 84% in round 3. |
| Mirza (76) | Multi-dimensional | Somalia | Multiple | Program evaluation | Complex humanitarian crisis. Multi-dimensional intervention implemented in setting of ongoing conflict and population displacement, with extensive damage to infrastructure (including health facilities). In this context, Child Health Days (CHDs) have been used regularly in Somalia to provide outreach health services to children <5. | Adaptive | | | | | | | | | | | Reducing cost and other access barriers to health services by bringing them to communities in Somalia. CHDs focused on delivery of an integrated service packaged including immunization (especially against measles and tetanus), vitamin A supplementation, oral rehydration therapy, use of insecticide-treated nets, and treatment of malaria. Mechanisms for improving uptake included the use of social mobilisation communities drawing in local leaders, and health worker training in logistics and monitoring. Wider system strengthening included use of CHD resources to support cold chain improvements that would last beyond the life-cycle of the intervention. | Analysis of health facility data and reported CHD coverage demonstrated that the proportion of regions with routine measles coverage >50% increased from 10% at baseline (using regular EPI activities only) to >84% in 2009 following the introduction of CHDs alongside regular EPI activities. |
| Ngwa (30) | Campaign | Nigeria | Cholera | Cross-sectional survey | Complex humanitarian crisis, driven primarily by conflict. The study focuses on campaign activities in Borno State in northern Nigeria in the context of an ongoing insurgency, population displacement, and poor health service access especially in outlying areas. | Adaptive | | | | | | | | | | | See above | 90% (95%CI: 88-92%) of the population targeted by the campaign received at least one dose of OCV. Weighted complete coverage was 73% (68-77%) among the target population. The highest increase in coverage between rounds was seen among girls aged 1-4 years. |

| | | | | | | *Mechanism* | | | | | | | | | | | | |
|---|---|---|---|---|---|---|---|---|---|---|---|---|---|---|---|---|---|---|
| | | | | | | | *Activities* | | | | | | | | | | | |
| | | | | | | | *System "hardware"* ⟷ *System "software"* | | | | | | | | | | | |
| ***First author*** | ***Intervention class*** | ***Country*** | ***Antigen*** | ***Study design*** | ***Context*** | *Type* | *Increased material resource availability* | *Increased human resource availability and capability* | *Changes to collaterals and redundancy* | *Changes to networks and collaboration* | *Increased flexibility (including decentralisation)* | *Improved situational awareness and information management* | *Improved preparedness and planning* | *Strengthened leadership* | *Changes to organisational and wider system culture* | *Improvement population trust* | *Description* | ***Outcome(s)*** |
| Ngwa (31) | Campaign | Nigeria | Cholera | Qualitative study | Complex humanitarian crisis, driven primarily by conflict. The study focuses on campaign activities in Borno State in northern Nigeria in the context of an ongoing insurgency, population displacement, and poor health service access especially in outlying areas. | Adaptive | | | | | | | | | | | Reactive OCV campaign. A key success factor identified was the fact that the Nigeria CDC had held a cholera preparedness workshop earlier in the year, including activities to lay the groundwork for an application to the global OCV stockpile, and agreeing mechanisms for deployment within-country. This was accompanied by provisional licensing for OCV in Nigeria which sped up necessary approvals when the outbreak began. Service delivery mechanisms drew on existing GPEI infrastructure, and polio microplanning templates originally developed in Sierra Leone. Comprehensive community engagement work was also carried out including segmenting messages by population groups and translating materials into local languages. | Reports a case fatality rate by the end of the outbreak of 1.14% - but this is the counterpart study to Ngwa (30) (see further results below) |
| Nkwogu (61) | Governance and coordination | Nigeria | Polio | Time series analysis | Conflict-affected setting. Focus on Borno State in northern Nigeria, in context of ongoing insecurity linked to insurgency, population displacement and constrained health service access especially in outlying areas. | Adaptive | | | | | | | | | | | Intervention focused on promoting civil-military engagement, through deployment of a civilian joint task force (CJTF), described as a "community-initiated security network" supporting the Nigerian military against the Boko Haram insurgency in the north of the country. CJTF members do not necessarily have military training but are drawn from the communities they are working with. CJTF brokering helped to secure agreements with local actors to permit "hit and run" vaccination activities during periods of active conflict, and to support movement of consumables and in-kind incentives to households to support uptake. | The principal effect of the intervention was to improve accessibility and led to a 47% increase in the number of wards perceived as accessible for vaccination staff over the 6 months of the intervention. The percentage of zero-dose children decreased from 8% to 3%. |

| First author | Intervention class | Country | Antigen | Study design | Context | Mechanism | | | | | | | | | | | | Outcome(s) |
|---|---|---|---|---|---|---|---|---|---|---|---|---|---|---|---|---|---|---|
| | | | | | | | Activities: System "hardware" ↔ System "software" | | | | | | | | | | | |
| | | | | | | Type | Increased material resource availability | Increased human resource availability and capability | Changes to collaterals and redundancy | Changes to networks and collaboration | Increased flexibility (including decentralisation) | Improved situational awareness and information management | Improved preparedness and planning | Strengthened leadership | Changes to organisational and wider system culture | Improvement population trust | Description | |
| Oladeji (56) | Service integration | South Sudan | Multiple | Time series analysis | Complex humanitarian crisis. Intervention implementation in South Sudan in context of ongoing conflict, population movement and high levels of underlying population vulnerability including malnutrition, limited health service access. The specific geographical focus of the intervention was a large IDP camp (Bentiu). | Adaptive | | | | | | | | | | | Integration intervention linking outpatient therapeutic program (OTP) centres (for malnutrition interventions) with neighbouring primary healthcare centres where immunisations are typically delivered. Children attended the OTP centres were screened for immunisation status and had any missing vaccinations administered by EPI staff attached to the centres for the duration of the intervention. Funds to support integrated delivery were pooled from nutrition and EPI sources. | The number of children vaccinated across all antigen types reported to have risen a year on from baseline analysis, but coverage estimates (and statistical significance) is not reported. Vaccination dropout rates were assessed to be lower from the OTP centres than from primary healthcare centres in the comparison arm (OR for dropout from OTP was 0.27 in one sector and 0.45 in another – both statistically significant). |
| Ongwae (55) | Service integration | Nigeria | Polio | Program evaluation | Conflict-affected setting. Focus of the intervention on six northern Nigeria states including Borno, a site of insecurity linked to insurgency at this time. Wider contextual features of the operating context in Borno are as described above for other studies based in this region. | Adaptive | | | | | | | | | | | Service delivery supported by MHTs and VCMs to target communities deemed at high risk for polio transmission. Primary mechanism for improving uptake was community engagement: communities were involved in selection of candidate VCMs who were then trained in vaccine administration, surveillance data gathering and other activities by implementing partner organisations. Traditional religious and community leaders were also engaged in selection of timing and approach to vaccination outreach activities. MHTs were used to increase reach in remote areas. | The percentage of children aged 12–23 months who were fully immunized rose from from 19% to 55% over the course of the intervention. |
| Oteri (65) | Health information and surveillance | Nigeria | Measles | Program evaluation | Conflict-affected setting. The intervention was national but this study includes focused discussion of challenges in implementation in Borno State – a context affected by insurgency-related insecurity as noted for other studies above. | Adaptive | | | | | | | | | | | Use of geographical information system (GIS) tools to support denominator estimation for eligible populations for a measles vaccination campaign. Comparison was made between accuracy of GIS-based estimates for northern states including Borno, and a walk-through methodology applied in southern Nigeria. GIS was used to map optimal post locations to ensure that no population settlement was more than 1km away from a site capable of delivering vaccinations – on the assumption that distance was a key barrier to access. | Mean variance of denominator estimates using GIS for northern states was lower (8.2%) than for southern states where the walk-through methodology was used (19.6%) – although these global estimates hid large variations by state and variation for Borno was particularly high (44.5%). Reasons for this are not clearly described in the study but could include ongoing population displacement after GIS estimates were generated. |

| | | | | | | Mechanism | | | | | | | | | | | | |
|---|---|---|---|---|---|---|---|---|---|---|---|---|---|---|---|---|---|---|
| | | | | | | | Activities: System "hardware" ↔ System "software" | | | | | | | | | | | |
| ***First author*** | ***Intervention class*** | ***Country*** | ***Antigen*** | ***Study design*** | ***Context*** | *Type* | *Increased material resource availability* | *Increased human resource availability and capability* | *Changes to collaterals and redundancy* | *Changes to networks and collaboration* | *Increased flexibility (including decentralisation)* | *Improved situational awareness and information management* | *Improved preparedness and planning* | *Strengthened leadership* | *Changes to organisational and wider system culture* | *Improvement population trust* | *Description* | ***Outcome(s)*** |
| Peyraud (37) | Campaign | Central African Republic | Multiple | Cross-sectional survey | Conflict-affected setting. Study describes a post-conflict vaccination in CAR, which had been in active conflict from 2012-15. Authors significant health service delivery challenges even before this, with only 48% of primary healthcare facilities having sufficient capacity to deliver EPI vaccines. Conflict-related damage to infrastructure was significant, and included outright destruction of around 25% of health facilities, and plundering of cold chain and logistics equipment. | Adaptive | | | | | | | | | | | Multi-pronged, multi-antigen campaign designed to provide a new baseline for childhood vaccination coverage, and targeting only children aged <1 in a single prefecture (state). The campaign was delivered as a partnership between the CAR Ministry of Health and MSF. Service delivery was integrated with other interventions dependent on the campaign round: distribution of vitamin A in the first round, nutritional screening in the second round and soap provision in the second and third rounds. | Statistically significant increases in coverage of all included antigens from pre- to post-campaign except for dose 3 PCV and the yellow fever vaccination (which was not introduced until the third round of the campaign). |
| Porta (44) | Campaign | South Sudan | Cholera | Program evaluation | Complex humanitarian crisis. Intervention implementation in South Sudan in context of ongoing conflict, population movement and high levels of underlying population vulnerability including malnutrition, limited health service access. Prior to the intervention, immunisation services had predominantly been delivered in a vertical way linked to specific programme funding. | Adaptive | | | | | | | | | | | Preventive mass vaccination campaign for cholera focused on four refugee camps and the surrounding host community population (on the basis of presumed contact between them and refugees e.g. in market settings). Campaign was carried out in two rounds. All people aged >1 yr were included, without intercurrent illness. Service delivery through a combination of fixed site and mobile outreach, where fixed sites were agreed with communities in advance, and outreach activities were designed to focus on common access points e.g. markets. Social mobilisation activities started around a week before the campaign to increase engagement. | Administrative coverage after round 1 was recorded as 84% or more (with some variations by geography) and 53-90% variably for dose 2 depending on the area. Intensity of vaccine administration was greater lower with the fixed site strategy (circa 250 vaccinations per day) whereas the outreach approach vaccinated (500-700 individuals per day). |
| Rainey (66) | Health information and surveillance | Haiti | Multiple | Program evaluation | Natural disaster – on background of a severe earthquake that struck Haiti in January 2010. This led to the emergence of temporary camps housing an estimated 1.5 million internally displaced children and adults. The wider health system context is described as fragile with long-standing resource shortages (financial and human) and variations in vaccination coverage by antigen, as well as nearer-term effects linked to destruction of facilities in the earthquake. | Adaptive | | | | | | | | | | | Focused on information gathering around a multi-antigen campaign administering either DTP or tetanus and diphtheria vaccine because a diphtheria outbreak had occurred in Haiti in 2009. A rapid monitoring approach was used to assess uptake following rounds of the campaign – using outreach workers to visit camps and settlements and effectively generate tallies for eligible children who were vaccinated and unvaccinated. | 23% of 310 vaccinated camps included in the study had been monitored by the time it concluded. 32 (44%) of those monitored were identified as targets for mop-up vaccination – a much higher than expected fraction likely because of ongoing population movement. However, only six of them had received repeat vaccination when further checks were carried out 6 weeks later. This marginal effect was likely due to lack of capacity among vaccinating teams to respond to need. |

| | | | | | | Mechanism | | | | | | | | | | | | |
|---|---|---|---|---|---|---|---|---|---|---|---|---|---|---|---|---|---|---|
| | | | | | | | Activities: System "hardware" ↔ System "software" | | | | | | | | | | | |
| **First author** | **Intervention class** | **Country** | **Antigen** | **Study design** | **Context** | *Type* | *Increased material resource availability* | *Increased human resource availability and capability* | *Changes to collaterals and redundancy* | *Changes to networks and collaboration* | *Increased flexibility (including decentralisation)* | *Improved situational awareness and information management* | *Improved preparedness and planning* | *Strengthened leadership* | *Changes to organisational and wider system culture* | *Improvement population trust* | *Description* | **Outcome(s)** |
| Rouzier (79) | Multi-dimensional | Haiti | Cholera | Program evaluation | Natural disaster – cholera outbreak following the 2010 earthquake, leading to an estimated 650,258 cases and 8,048 as of March 2013 (the time at which the study was conducted). Wider health system context of fragility including limited access to clean water (63% of the population has access to improved water) and sanitation (17% access to improved sanitation), as well as high levels of population need (ongoing epidemics of HIV and tuberculosis). | Adaptive | | | | | | | | | | | Multi-dimensional intervention to deliver cholera vaccination in proof-of-concept intervention targeting residents of urban slums in the Haitian capital. Intervention drew on established position of a NGO serving this population, which set up a new central coordinating committee, novel communication plans, and engagement with community leaders to bolster uptake for OCV. Large investments were made in human resourcing, for which 20% of the total operating staff of the NGO were transferred to support the intervention, and training in vaccine administration and monitoring. | 90.8% of the population identified as eligible received two OCV doses, with an estimated overall vaccine coverage among the local population of 74.8%. Dose wastage was very low – 99.9% of doses received by the NGO for the campaign were reported as administered. |
| Sheikh (42) | Campaign | Kenya | Polio | Program evaluation | Complex humanitarian crisis. Reactive campaign in response to seven polio cases in refugee camp residents and six in surrounding communities near the Kenya-Somalia border. Broader context of large, semi-settled refugee population in Kenyan border regions but ongoing displacement linked to conflict in Somalia. | Adaptive | | | | | | | | | | | Reactive campaign. Under leadership of the Kenyan Ministry of Health, one national and five subnational OPV campaigns were carried out between May–November 2013, followed by a combined IPV-OPV combined in a campaign directed at approximately 126,000 children aged ≤59 months including those who lived in five refugee camps and surrounding communities near the Kenya-Somalia border. Service delivery was through a combination of fixed sites and mobile outreach and was accompanied by community mobilisation activities informed by lesson learning from the GPEI in Kenya. | Coverage (based on caregiver recall) with OPV and IPV in the December campaign was 92.8% in the refugee camps and 95.8% in surrounding communities. Receipt of OPV in the November campaign was 97.2% in the refugee camps and 97.3% in surrounding communities. |
| Shuaibu (32) | Campaign | Nigeria | Polio | Program evaluation | Conflict-affected setting. Focus on Yobe and Borno States which reported 46.3% of all polio cases in Nigeria during the study period, but are affected by significant security challenges linked to ongoing insurgency. Wider health system context that some areas of these states are inaccessible, and surveillance is an ongoing challenge. Armed conflict has contributed to destruction of health facilities and cold chain equipment. | Adaptive | | | | | | | | | | | Reactive vaccination campaign involving introduction of IPV alongside three-dose OPV for polio, coordinated through both national and state-level polio emergency control centres. Service delivery was supported by temporary recruitment of vaccinators from the private and voluntary sectors alongside public providers, followed by training in vaccination administration and monitoring. Community mobilisation was supported by targeted messaging and use of community roundtables among other routes. | Administrative coverage in Yobe (OPV) and Borno (IPV) was in excess of 100% following the campaign – suggesting challenges in denominator estimation in the face of ongoing population displacement. IPV coverage in Yobe was 94.9% following the campaign (again, administrative). |

| | | | | | | Mechanism | | | | | | | | | | | | |
|---|---|---|---|---|---|---|---|---|---|---|---|---|---|---|---|---|---|---|
| | | | | | | | Activities: System "hardware" ↔ System "software" | | | | | | | | | | | |
| ***First author*** | ***Intervention class*** | ***Country*** | ***Antigen*** | ***Study design*** | ***Context*** | *Type* | *Increased material resource availability* | *Increased human resource availability and capability* | *Changes to collaterals and redundancy* | *Changes to networks and collaboration* | *Increased flexibility (including decentralisation)* | *Improved situational awareness and information management* | *Improved preparedness and planning* | *Strengthened leadership* | *Changes to organisational and wider system culture* | *Improvement population trust* | *Description* | ***Outcome(s)*** |
| UNICEF Lebanon (77) | Multi-dimensional | Lebanon | Multiple | Program evaluation | Complex humanitarian crisis, featuring large-scale population influx from Syria and deteriorating economic conditions among others. Wider health system context of provider fragmentation for childhood vaccination, including a prominent role for the private sectors, although refugees and host communities alike are theoretically able to access EPI vaccination free of charge through publicly-supported providers. | Adaptive | | | | | | | | | | | SIAs led by Lebanese Ministry of Health and UNICEF, with implementing partner support. Multiple service delivery modes but emphasis on publicly-supported primary healthcare facilities, and dispensaries (pharmacies) supported by the Ministry of Health. Outreach activities were also extensively used to identify zero-dose or undervaccinated children and refer in to fixed sites for vaccine administration. | National coverage estimation for the study period was not available at the time the report was written. However, findings show a progressive increase in the number of zero-dose children identified at each SIA round, from 65% in round 1 (of whom 44% were then vaccinated); 78% during round 2 (51% vaccinated); and 85% in round 3 (55% later vaccinated). |
| Usman (68) | Community engagement and mobilisation | Nigeria | Polio | Program evaluation | Conflict-affected setting. Implementation in northern states in Nigeria including Borno (see above for references to impact of Boko Haram insurgency). Wider health system setting in which parties to the conflict had targeted and killed polio workers, disrupted polio campaigns, and damaged or destroyed health facilities. | Adaptive | | | | | | | | | | | Focusing on expanded community engagement and mobilisation, with a particular focus on tackling perceived misinformation regarding polio vaccination. Intervention implemented with central coordination from the national emergency operations centre for polio. VCMs recruited under the programme are women drawn from the communities they are working with, and work using intensive outreach. Further efforts to strengthen trust by working in partnership with community and religious leaders, among other strategies. | First dose OPV coverage increased from 54.8% in 2014 to 99.0% in 2018 and OPV3 coverage among children aged 12 – 23 months increased from 47.2% in 2014 to 88.4% in 2018. The percentage of fully immunized (all antigens) children rose from 33.0% at baseline in 2014 to 67.0% in 2018, illustrating spillover effects on trust in vaccination in general arising from a community mobilisation effort focused primarily on polio. |
| Valadez (48) | Health financing | South Sudan | Multiple | Program evaluation | Complex humanitarian crisis. Intervention implementation in South Sudan in context of ongoing conflict, population movement and high levels of underlying population vulnerability including malnutrition, limited health service access. Prior to the intervention, immunisation services had predominantly been delivered in a vertical way linked to specific programme funding. | Adaptive | | | | | | | | | | | Macro-level financing through donors to support health system strengthening in South Sudan in the context of ongoing conflict. Targets for financing included improving human resource capacity at the Ministry of Health, health financing, governance and information systems, and strengthening service delivery and informatics. World Bank financial support specifically addressed delivery of a basic health services package including childhood immunisation. Service delivery changes linked to funding included task-shifting and health worker capacity building/training. | Measles vaccination coverage in children aged 12–23 months increased by 11.2% (p<0.001) to 49.7%. Child immunisation coverage overall improved significantly by the study end-point (p<0.001) but remained low with about half of the children obtaining a measles vaccination (49.7%, 95% CI ±2.8), and 20.8% in receipt of all childhood immunisations (±2.3%). |

| | | | | | | Mechanism | Activities: System "hardware" ↔ System "software" | | | | | | | | | | | |
|---|---|---|---|---|---|---|---|---|---|---|---|---|---|---|---|---|---|---|
| ***First author*** | ***Intervention class*** | ***Country*** | ***Antigen*** | ***Study design*** | ***Context*** | *Type* | *Increased material resource availability* | *Increased human resource availability and capability* | *Changes to collaterals and redundancy* | *Changes to networks and collaboration* | *Increased flexibility (including decentralisation)* | *Improved situational awareness and information management* | *Improved preparedness and planning* | *Strengthened leadership* | *Changes to organisational and wider system culture* | *Improvement population trust* | *Description* | ***Outcome(s)*** |
| Venczel (43) | Campaign | Haiti | Measles | Program evaluation | Complex humanitarian crisis. Broader context in Haiti of high levels of inequality, poverty and globally poor health outcome indicators including low life expectancy and high rates of HIV and tuberculosis. Wider health system context including limited health service access especially for low-income urban populations, and low resourcing for health services in general. | Adaptive | | | | | | | | | | | Reactive vaccination campaign in response to emergent measles outbreak, incorporating door-to-door outreach and vaccination, using cadres of vaccinators some of whom were recruited and trained specifically for the campaign. Uptake was independently monitored in parallel to the campaign. The authors note that ability to carry out the campaign depended on mobilisation of significant financial resources almost all of which came from outside Haiti. | Post-campaign monitoring activities indicated coverage in the eligible population (children <10) of 88% but with significant variations in coverage across geographical areas targeted. |
| Vink (49) | Health financing | Afghanistan | Multiple | Cross-sectional survey | Conflict-affected setting. Wider context in Afghanistan at the time of the study has been described for other included studies above. This intervention focused on a single province – Uruzgan – where 97% of the population lives in rural areas, and health outcomes are poor: 30.4% of pregnant women make use of antenatal services and during their delivery only 23.5% are assisted by a skilled birth attendant; 5.7% of the children aged 12-23 months received all necessary vaccinations prior to the intervention. | Transfor-mative | | | | | | | | | | | Public-private partnership between Dutch NGO and private providers in Uruzgan, drawing in 34 private providers to administer childhood vaccinations. This was achieved through introduction of financial incentives to replace lost income for private providers, and was supported by training for health providers in the partnership on vaccination techniques, safety/hygiene methods and monitoring. Facilities were renovated and consumables supplied. A local council for each clinic was also set up to strengthen relations with the community. | DTP3 and MCV1 coverage were significantly higher in intervention villages (73.5%, 66.4% and 69.9% respectively) than in control villages (36.0%, 5.2% and 26.2% respectively; P < 0.0001 for all comparisons). The proportion of children being fully vaccinated (excluding BCG) was 54.9% in the PHP villages and 4.9% in the control villages (P < 0.0001). A key shift was in the location of vaccination – in control areas, most children were vaccinated in mass campaigns, whereas most in intervention areas were vaccinated at fixed sites. |
| Warigon (69) | Community engagement and mobilisation | Nigeria | Polio | Program evaluation | Conflict-affected setting. Focus on northern Nigerian states including Borno and Yobe (conditions in these areas are as described for other studies included above). | Adaptive | | | | | | | | | | | Increasing uptake through community mobilisation and engagement. These included (i) use of in-kind incentives to increase uptake (e.g. foodstuffs); (ii) road shows; (iii) engagement through local religious leaders; (iv) engagement with nomadic communities through nominated intermediaries; (v) outreach into security-compromised zones, and (vi) health camps. All of these activities were focused on improving trust in vaccine providers, and addressing reasons for hesitancy, to increase demand. | The proportion of children with non–polio-associated AFP who received at least four OPV doses rose from 80% in 2012 to 97% in 2014. The proportion of children who were underimmunized – defined as incomplete OPV courses – declined from 17% to 4%. The proportion of zero dose children also declined from 3% to 1% over the time course of the intervention. |

| | | | | | | Mechanism | | | | | | | | | | | | |
|---|---|---|---|---|---|---|---|---|---|---|---|---|---|---|---|---|---|---|
| | | | | | | | Activities<br>System "hardware" ↔ System "software" | | | | | | | | | | | |
| ***First author*** | ***Intervention class*** | ***Country*** | ***Antigen*** | ***Study design*** | ***Context*** | *Type* | *Increased material resource availability* | *Increased human resource availability and capability* | *Changes to collaterals and redundancy* | *Changes to networks and collaboration* | *Increased flexibility (including decentralisation)* | *Improved situational awareness and information management* | *Improved preparedness and planning* | *Strengthened leadership* | *Changes to organisational and wider system culture* | *Improvement population trust* | *Description* | ***Outcome(s)*** |
| WHO/GPEI (45) | Campaign | Multiple | Polio | Other | Multiple sites addressed, but the two of immediate relevance to this study were Somalia (complex humanitarian crisis featuring chronic insecurity with limited access in several areas of the country, and ongoing cross-border and within-border population movement); and Pakistan, particularly in the context of ongoing conflict following military operations, where Swat valley (the focus of the GPEI intervention discussed) was inaccessible between November 2008 and August 2009. | Adaptive | | | | | | | | | | | Change in dosing strategy for OPV, and the implications this has for service delivery. Shorter interval dosing allowed concentration of field supervisors over a shorter time period to provide stronger oversight of vaccination delivery across populations in both settings, but also depended on stronger community engagement and advocacy activities to sensitive communities to the likelihood of multiple dose administration in a campaign context. | For the two case study countries: the shorter interval dosing campaign in Somalia resulted in an early end to polio transmission in these areas by comparison with control areas. In Pakistan, administrative coverage following the campaign was 93%, and 86% by post-campaign monitoring, suggesting high uptake is possible given a higher intensity campaign. |
| Yehuala-shet (64) | Health workforce | Nigeria | Polio | Program evaluation | Conflict-affected setting. Intervention implemented across northern Nigerian states, but particularly including conflict-affected States in the north of the country such as Borno and Yobe (see context notes for other studies above). | Adaptive | | | | | | | | | | | Recruitment and deployment of technical surge capacity to support improved polio vaccine uptake in the context of ongoing transmission particularly in the north of the country. WHO worked with the Ministry of Health and other domestic actors to identify a roster of technical experts, and risk-assess geographical regions for the extent of transmission. These inputs were used to assign (on a biannual basis) technical experts to underperforming areas, so that the most experienced experts were allocated to areas were current performance was poorest. The intervention was underpinned by use of variable contract types. | Substantial improvements noted in core polio prevention indicators (e.g. non-polio AFP rate, and stool sample adequacy). The proportion of wards across these areas with >10% children missed by vaccination campaigns decreased from 21% in 2012 to 3% in 2015. |

| | | | | | | Mechanism | | | | | | | | | | | | | |
|---|---|---|---|---|---|---|---|---|---|---|---|---|---|---|---|---|---|---|---|
| | | | | | | | Activities: System "hardware" ⟷ System "software" | | | | | | | | | | | | |
| ***First author*** | ***Intervention class*** | ***Country*** | ***Antigen*** | ***Study design*** | ***Context*** | *Type* | *Increased material resource availability* | *Increased human resource availability and capability* | *Changes to collaterals and redundancy* | *Changes to networks and collaboration* | *Increased flexibility (including decentralisation)* | *Improved situational awareness and information management* | *Improved preparedness and planning* | *Strengthened leadership* | *Changes to organisational and wider system culture* | *Improvement population trust* | *Description* | | ***Outcome(s)*** |
| Yehuala-shet (53) | Health financing | Nigeria | Multiple | Program evaluation | Conflict-affected setting. Intervention implemented nationally, but included conflict-affected States in the north of the country such as Borno and Yobe (see context notes for other studies above). | Adaptive | | | | | | | | | | | Introduction of direct payment disbursement mechanism to ensure timely payment of eligible vaccination personnel, improve payment transparency, and release operational funds quickly to local areas to support vaccination delivery. WHO and UNICEF worked with the Nigerian government and with a leading bank to set up disbursement systems, but also established 280 new payment points in less accessible parts of the country to speed up access to funds for health workers operating in these areas. Guidance on budgets and spending allocations were developed and distributed by WHO prior to each round. The aim of the intervention was to promote vaccination delivery through timely financing of local vaccination teams. | | Total volume of funds disbursed via this mechanism increased year on year following its introduction. Percentage variation between funds disbursed and accounted for for SIA activities declined to 0% within three years of the introduction of the intervention. |
